# Ondansetron use is associated with lower COVID-19 mortality in a Real-World Data network-based analysis

**DOI:** 10.1101/2021.10.05.21264578

**Authors:** Gregory M. Miller, J. Austin Ellis, Rangaprasad Sarangarajan, Amay Parikh, Leonardo O. Rodrigues, Can Bruce, Nischal Mahaveer Chand, Steven R. Smith, Kris Richardson, Raymond Vazquez, Michael A. Kiebish, Chandran Haneesh, Elder Granger, Judy Holtz, Jacob Hinkle, Niven R. Narain, Bret Goodpaster, Jeremy C. Smith, Daniel S. Lupu

## Abstract

**Objective:** The COVID-19 pandemic generated a massive amount of clinical data, which potentially holds yet undiscovered answers related to COVID-19 morbidity, mortality, long term effects, and therapeutic solutions. The objective of this study was to generate insights on COVID-19 mortality-associated factors and identify potential new therapeutic options for COVID-19 patients by employing artificial intelligence analytics on real-world data.

**Methods:** A Bayesian statistics-based artificial intelligence data analytics tool (bAIcis®) within Interrogative Biology® platform was used for network learning, inference causality and hypothesis generation to analyze 16,277 PCR positive patients from a database of 279,281 inpatients and outpatients tested for SARS-CoV-2 infection by antigen, antibody, or PCR methods during the first pandemic year in Central Florida. This approach generated causal networks that enabled unbiased identification of significant predictors of mortality for specific COVID-19 patient populations. These findings were validated by logistic regression, regression by least absolute shrinkage and selection operator, and bootstrapping.

**Results:** We found that in the SARS-CoV-2 PCR positive patient cohort, early use of the antiemetic agent ondansetron was associated with increased survival in mechanically ventilated patients.

**Conclusions:** The results demonstrate how real world COVID-19 focused data analysis using artificial intelligence can generate valid insights that could possibly support clinical decision-making and minimize the future loss of lives and resources.

## INTRODUCTION

### Background and significance

As of September 2021, an estimated 221 million cases of COVID-19 and over 4.5 million deaths have been reported worldwide [1]. The United States has reported the largest proportion of COVID-19 cases estimated at 40 million with approximately 650,000 reported deaths [1]. Worldwide efforts are currently focused on the implementation of an aggressive vaccination program to control the pandemic. Despite the control strategies of limiting COVID-19 infections by physical measures (use of masks, isolation, social distancing) and vaccination, the emergence of new SARS-CoV-2 variants across the globe, the increased incidence of breakthrough infections, especially in the younger population, and the evolving understanding of infection cycles and re-emergence provides impetus for continued investigation of real-world data (RWD). This investigation can generate insights into disease susceptibility and long-term effects, and as well can provide potential therapeutic strategies.

The revolution in computational analytics, including the considerable progress achieved in the application of artificial intelligence (AI) and machine learning (ML) capabilities, in tandem with access to high-density RWD and clinical evidence, provides a suitable environment to generate hypothesis-agnostic insights for the management of health and disease. Further, the availability of supercomputers and cloud-based high-performance computing capabilities significantly increases analytical depth and reduces time required to perform higher order AI/ML analytics of large population-based datasets, thus permitting a better understanding of disease etiology and facilitating the identification of novel information pertinent to disease management.

AI has been extensively applied to analyze various COVID-19 data, including to aid diagnostics and in therapeutic design [2]. In RWD AI, machine learning has been used to predict the probability of ARDS based on the clinical characteristics of COVID patients [3]. A further study, on 3,194 COVID-19 cases in the Emory Healthcare network, assessed whether a COVID-19 patient’s need for hospitalization can be predicted at the time of their RT-PCR test using electronic medical records data prior to the test [4].

Although concern has been raised about the use of untested AI programs and small data sets in COVID research [5], AI continues to play a major role in COVID decision making. Of particular importance in the current pandemic is finding causal relationships for disease outcomes, and in this respect, Bayesian networks are ideal for taking an event that occurred and predicting the likelihood that any one of several possible causes was the main contributing factor. Bayesian networks create a network of dependency links among variables of interest that enable inference of causality [6]. Such analysis has the benefit of determining which independent variables are directly associated with a clinical outcome variable of interest (e.g., death, admission to intensive care unit), and which variables are located farther in the chain of causality. A drawback of Bayesian networks is that their computational complexity is relatively high, but this can be overcome with sufficiently large computational power, as we did in our study.

The current study combines a large amount of COVID-19-focused RWD from AdventHealth that was analyzed using a Bayesian statistics driven platform on a supercomputer at Oak Ridge National Laboratory (ORNL). This collaboration allowed us to leverage the data availability and computational capacities in a data-driven manner to generate causal network models to identify factors associated with disease severity, increased survival and mortality, including drugs that have the potential to improve outcomes in COVID-19 patients. We used the bAIcis algorithm [7] to develop Bayesian networks based on various patient subpopulations and patient features from different time windows, before and after COVID-19 diagnosis, in order to identify those variables having a likely causal association with mortality. One of our key findings is that mortality has a dependency on the use of ondansetron, a widely used anti-emetic medication. After examining the interaction of ondansetron use with other variables of interest previously found to predict death, we found that the beneficial effect of ondansetron on mortality is specifically seen in patients on mechanical ventilator.

## MATERIALS AND METHODS

### Data collection

AdventHealth, headquartered in Orlando, FL, is one of the largest non-profit health care systems in the United States with over 50 hospitals in 12 states, and 5 million patient encounters annually (including inpatient, outpatient, and emergency visits). Early in the SARS-CoV-2 pandemic, AdventHealth established the Registry and Biorepository of COVID-19 (RECOVER-19), a registry of all patients tested for SARS-CoV-2 within the AdventHealth Enterprise (IRBnet # 1590483). The registry comprised raw data extracted from the AdventHealth Cerner Electronic Health Records (EHR) system and was made available in the Data Lake powered by the Integrated Data for Enterprise Analytics (IDEA) platform. The registry collected associated data from inpatients and outpatients, structured in 9 data tables, using the approach described in the supplemental methods (Supp. Table 1): Patient IDs, COVID-19 encounters, Diagnoses, Problems (patient personal medical history starting 2016), Procedures related to the COVID-19 visit, Clinical Events, Lab Results, In-house Medication Administration Records (MAR), Recorded Home and Prescription Discharge medications. To allow for greater user control, the Clinical Classifications Software Refined (CCSR) was implemented [8]. This study was approved by the AdventHealth Institutional Review Board (#1590483).

### Cohort selection

The RECOVER-19 registry, which is continuing to collect data, included 279,281 inpatients and outpatients tested for SARS-CoV-2 infection by antigen, antibody, or PCR methods from January to December 2020. From the 35,504 positive patients thus found, we selected for this analysis only the 16,277 found positive by PCR, due to higher confidence in this method (Figure 1).

**Figure 1.**
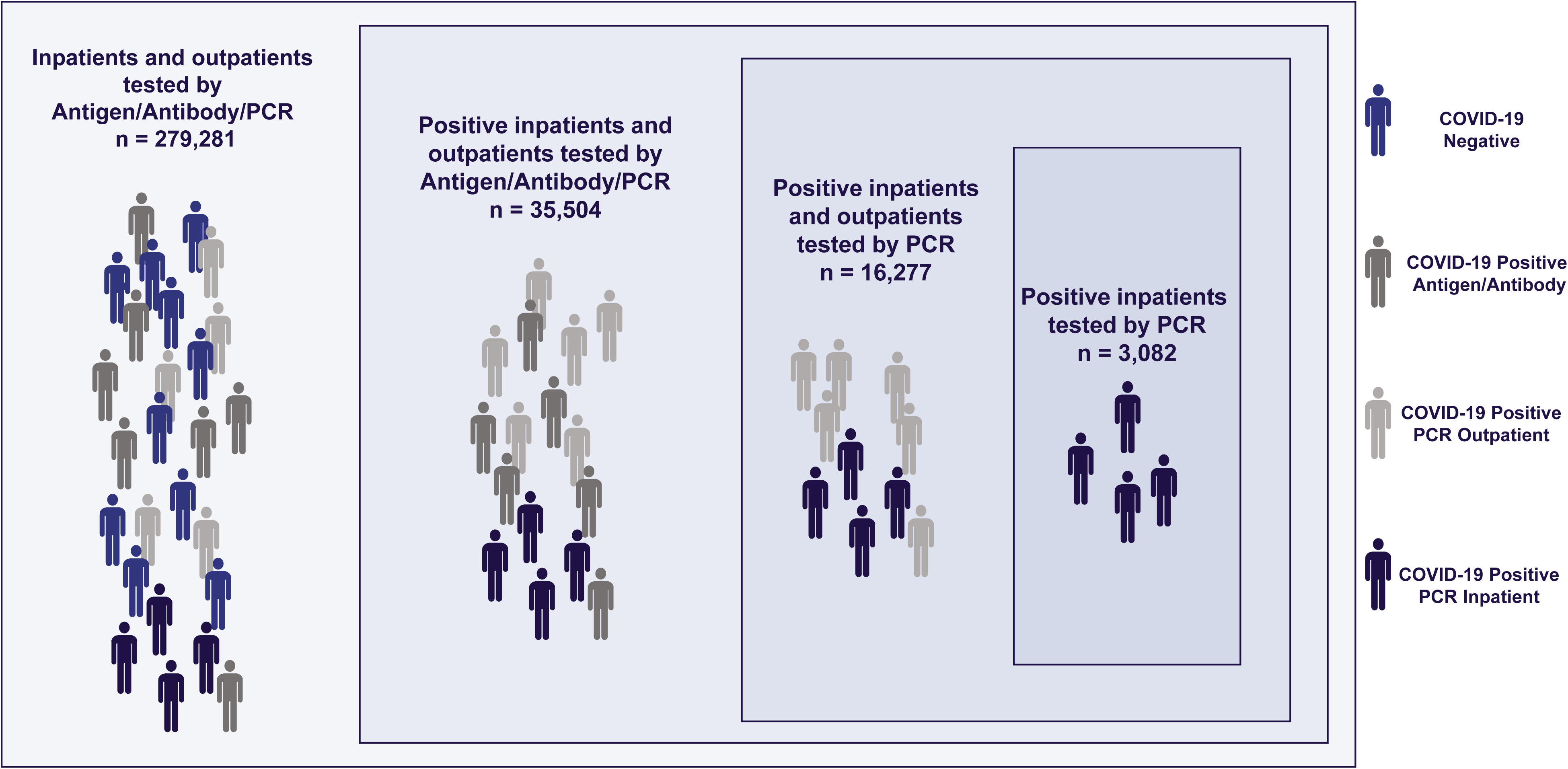
The RECOVER-19 registry included 279,281 inpatients and outpatients tested for SARS-CoV-2 infection by antigen, antibody, or PCR methods from January to December 2020. 16,277 PCR positive patients were selected for this analysis.

### Endpoint definitions

COVID-19 outcome variables (endpoints) derived in this project were: mortality, mechanical ventilation initiation (“being placed on a ventilator”), hospital admission (“being admitted to inpatient setting”), ICU admission (“being admitted to the ICU”), and length of stay (LOS). Specific definitions for endpoints can be found in the supplemental methods.

### Alignment of patient journeys

To enable Bayesian network inference, patients were aligned along their COVID-19 disease journey and features were defined in eleven time bins in relation to the time of COVID-19 positive PCR specimen collection. The time windows utilized in feature generation were: > 12, 9 - 12, 6 - 9, 3 - 6, 1 - 3, and < 1 *months prior* to the time of COVID-19 positive PCR test sample collection and < 7, 7 - 14, 14 - 21, 21 - 28, and > 28 *days after* the time of COVID-19 positive PCR test sample collection (Figure 2). Using these time bins, features were derived from all data tables.

**Figure 2.**
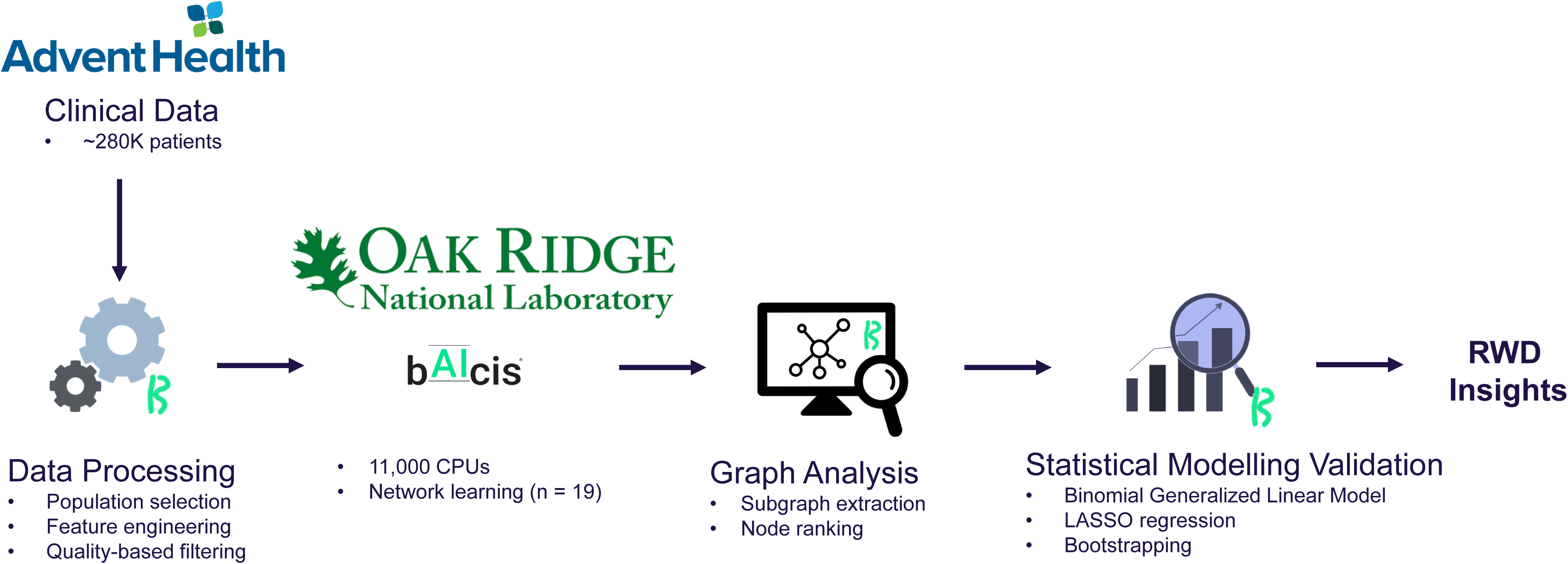
Overview of the data processing workflow.

### Bayesian network learning and hypotheses generation

BERG’s Bayesian AI analytics (bAIcis®) learning was employed to generate hypotheses of factors related to mortality. This unbiased approach yielded 19 networks representing the entire patient population as well as patient subpopulations (e.g., patients of similar age or ethnicity) (Table 1). Bayesian analytics is useful in generation of causal networks, in which nodes represent the analyzed features and edges the causal relationships, and where an upstream/parent node drives changes in downstream/child nodes. In this context, bAIcis® allows the pre-definition of causal hierarchies, so variables can be constrained to not have parent-nodes (constrained as ‘top’) or child-nodes (constrained as bottom), such as, for example, the top variable, “Age,” that is not driven by any other variable. In this regard, the top and bottom variables were selected in alignment with the data structure, and Bayesian networks were inferred containing the potential cause-and-effect relationships. Following bAIcis® learning, features related to mortality were identified by the following approach. First, subgraphs were extracted from the original bAIcis® networks by removing infrequent edges (edges present in the ensemble model with frequency ≤20%), then extracting the nodes connected to the mortality node by 1st, 2nd, or 3rd degree (Table 1, Column 4). All nodes selected by these criteria were then assessed by univariate statistical analysis to have a significant relationship to mortality in the respective patient cohort used for network learning. Features with p-value ≥ 0.05 by Fisher’s exact test or 0.67 ≥ odds ratio (OR) ≤ 1.5 were considered insignificant and disregarded in further analysis. Multiple testing correction was not applied to allow for more features to be included in downstream analysis.

**Table 1.**
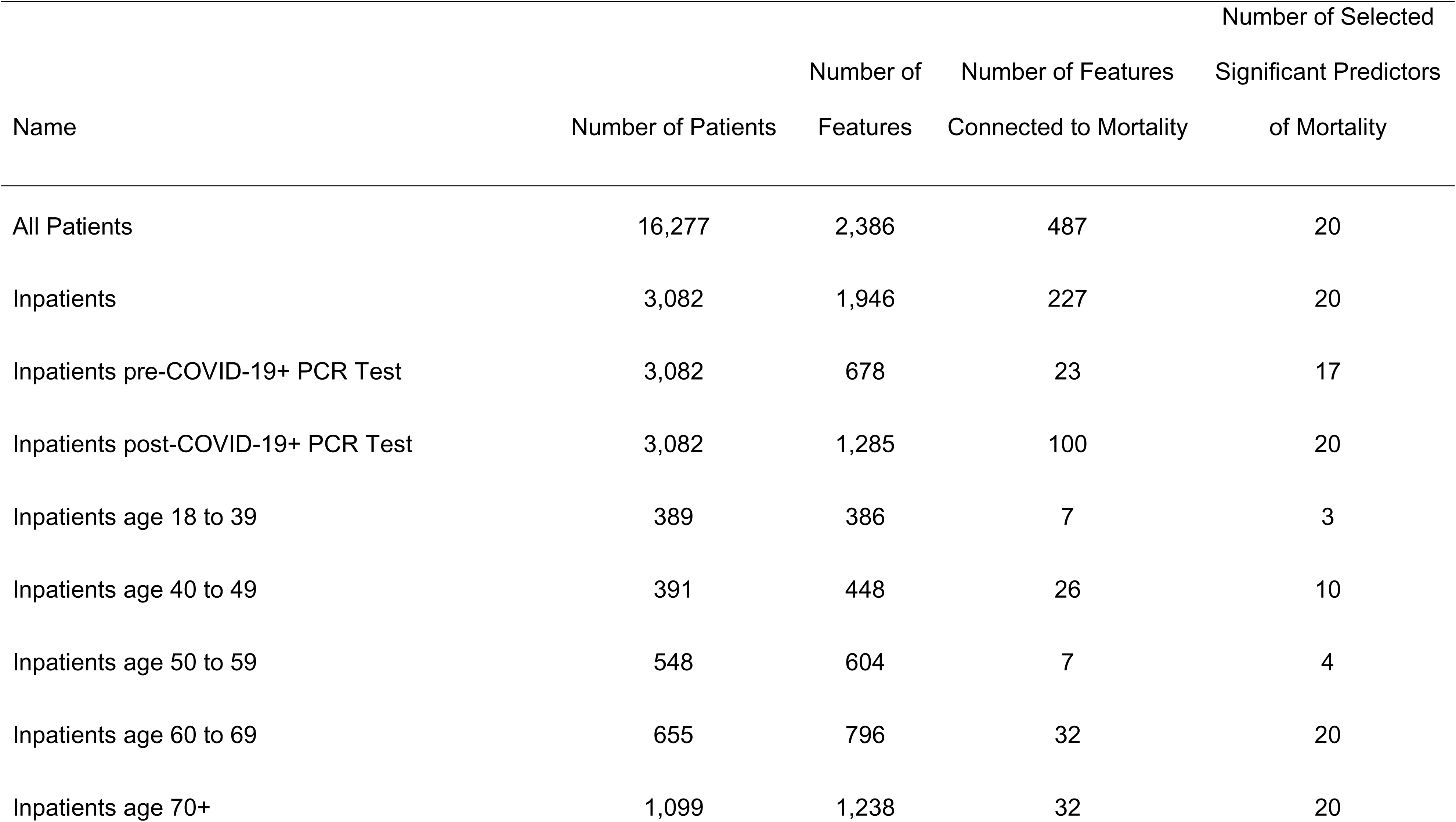
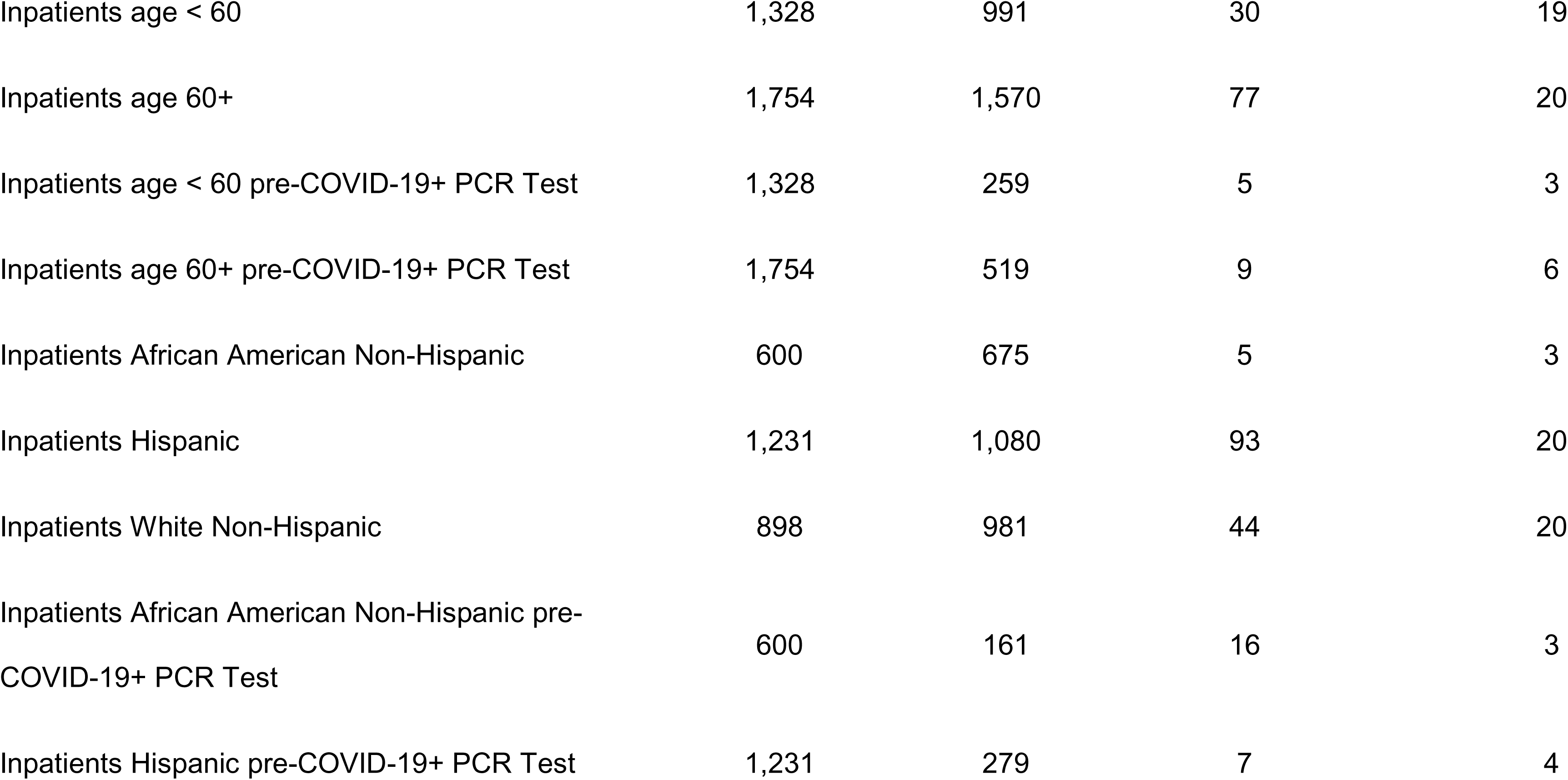
BERG’s Bayesian AI analytics (bAIcis*®*) generated 19 networks which enabled unbiased identification of significant predictors of mortality for specific patient populations.

### Computational resources for Bayesian network training

The deep Bayesian networks for this COVID-19 effort were trained using the Andes supercomputer at the Oak Ridge Leadership Computing Facility (OLCF). Andes is a 704-node machine with two AMD EPYC 7302 CPUs per node and is primarily focused on large-scale scientific discovery via data processing and modeling. The BERG bAIcis® software that depends on Apache Mesos was ported to run on Andes using Singularity containers. All models were trained with 32 Andes nodes and 8 Mesos tasks per node. Given the various dataset sizes (Table 1), Andes enabled a calculation of the Bayesian models with a median runtime of 178 seconds and a maximum runtime of 4.91 hours.

### Multivariable regression analysis for assessment of validity of ondansetron effect on COVID-19 associated mortality

A multivariate regression analysis was undertaken to determine the robustness of the Bayesian prediction of ondansetron discovery on mortality after adjusting for demographics and clinical variables (potential confounders). The following groups, comprising 25 variables, were examined using multi-variable regression for their ability to predict mortality in a COVID-19 positive patient cohort: Demographics (age (approximate), gender, race, and ethnicity), Medications (Ondansetron, Azithromycin, Remdesivir, Dexamethasone, Tocilizumab, Convalescent plasma), Comorbidities (Diabetes, Chronic obstructive pulmonary disease (COPD), Asthma, coronary artery disease (CAD), Heart Failure, Neoplastic Disease, Kidney Disease), Laboratory Analytes (C-reactive peptide (CRP), D-dimer, alanine amino transferase (ALT), Ferritin, aspartate transaminase (AST), blood urea nitrogen (BUN), Lymphocytes), and ventilator status. The de-identified dataset had patient ages structured in bins, such as 18 - 39, 40 - 49, …, 70+. For the purpose of regression modeling these were converted to approximate ages as 35 for the 18 - 39 bin, 45 for the 40 - 49 bin, and so on. Out of the 3,082 inpatients, 2,259 had complete observations for the variables indicated above. To increase the statistical power, imputation of missing values was accomplished by a multiple imputation approach using the predictive mean matching method [9], as implemented by the R package **mice** [10].

To study the ondansetron-associated effect of the above variables on COVID-19 mortality, a logistic regression was performed in a generalized linear model for the binary outcome of deceased or survived, using the R package **glm**. Subsequent analysis focused on fitting the logistic regression model to the 25 clinical variables (described above), the corresponding 300 (25 x 24/2) interaction terms (each equaling the pairwise product of their binary values) and the 8 squared terms for the continuous terms, for a total of 333 terms. Because of the inherent limitation of logistic regression in fitting such a model to the available data, we used the LASSO (least absolute shrinkage and selection operator) regression method (as implemented by the R package **glmnet** (6), with the parameter *alpha* set to 1) to select those covariates most likely to have non-zero coefficients. The *lambda* parameter was optimized with a cross-validation approach using the cv.glmnet function. *Lambda* was set conservatively to a value that is one standard deviation away from the minimum value determined from the cross-validation approach. Bootstrapping was performed by sampling with replacement of the dataset, while retaining the proportion of deceased to survived patients in the bootstrap datasets.

## RESULTS

### Cohort Selection for Discovery of Significant Predictors of COVID-19 Mortality

We stratified the RECOVER-19 registry patients based on SARS-CoV-2 Antigen, Antibody, and PCR test results, as well as inpatient vs outpatient status (Figure 1). The corresponding workflow of data processing, analysis for generation of causal network and predictive models is described in Figure 2.

### Features Associated with Increased COVID-19 Mortality

The workflow was initiated by reviewing data from 279,281 inpatients and outpatients tested for SARS-CoV-2 infection by antigen, antibody, or PCR methods from January to December 2020 in the AdventHealth Central Florida Division (CFD) health system. Data collected for those 16,277 PCR tested patients that were SARS-CoV-2 positive were utilized for the generation of causal networks using bAIcis® analysis network learning and hypothesis generation. We generated 19 networks which enabled unbiased identification of significant predictors of mortality at any time for specific patient populations (Table 1, Supp. Table 3). These networks included inpatient cohorts of multiple age groups, races, and ethnicities, before (pre-COVID) and after (post-COVID) their SARS-CoV-2 PCR positive test. The “All patients” causal network includes the full patient cohort of 16,277 and exhibited 2,386 features, 487 of which were connected to mortality, with 20 being significant predictors of mortality. Other networks with high numbers of significant features were “Inpatients age 60 to 69”, “Inpatients age 70+”, “Inpatients Hispanic”, and “Inpatients White Non-Hispanic”. Of the 239 significant mortality features identified in specific patient subpopulations, the majority (223/239, were found to be associated with increased mortality at any time (Supp. Table 2). Thirteen of the identified significant mortality features had more than two factor values (Supp. Table 4).

As expected, being placed on a ventilator or being admitted in the intensive care unit (ICU) was found to be associated with increased mortality consistently across 16/19 networks (Supp. Table 3) and 11/19 networks, respectively. Similarly, the length of stay was found to be associated with mortality (with longer stays associated with higher mortality) across 6/19 networks (Table 1). As expected, inpatient medications commonly administered in the ICU setting, such as fentanyl, midazolam, and cisatracurium were also found to be associated with increased mortality across multiple networks.

### Features Associated with Decreased COVID-19 Mortality

Three of the 239 significant mortality features were found to be associated with decreased mortality (Table 2), two of which referred to the use of ondansetron in patients younger than 60 years (Supp. Table 3). Within this patient population, in hospital ondansetron treatment either early (< 7 d) or late (> 28 d) after COVID-19 positive PCR test was found to decrease mortality (p = 0.03, OR = 0.45 and p = 0.001, OR = 0.079, respectively) (Figure 3). The third feature associated with decreased mortality was the use of ICD-10 code Z20.828 (“Contact with and (suspected) exposure to other viral communicable diseases”).

**Figure 3.**
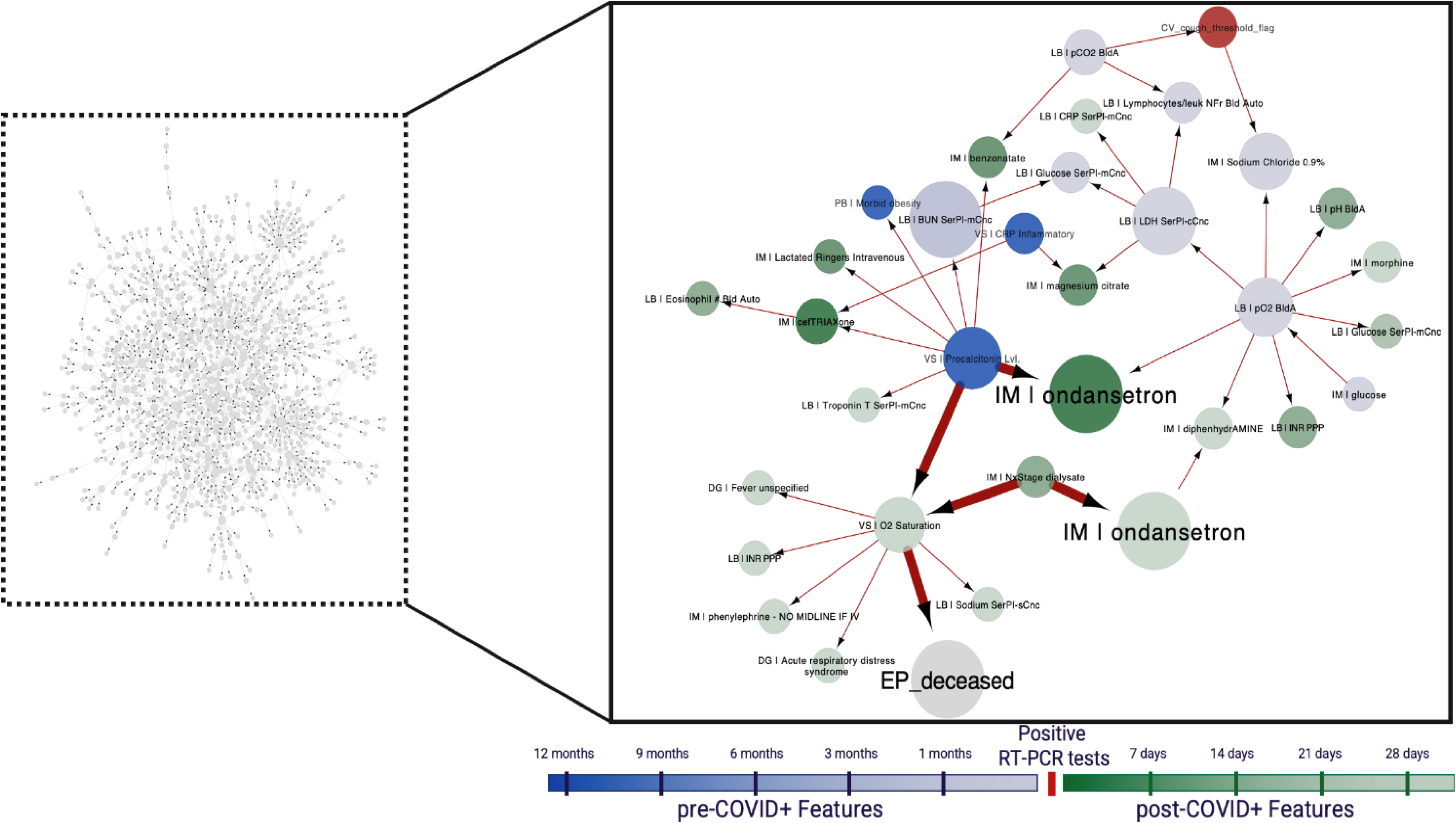
An illustrative example of the analysis approach. Left: A visualization of the bAIcis network learned from the age < 60 population admitted to the inpatient setting (n = 1,328). Right: Subgraph of the “Inpatients age < 60” bAIcis ® network illustrating the linkage between ondansetron use and mortality. Ondansetron use within the first week and after 28 days post-COVID-19 positive PCR test was significantly associated with decreased mortality (highlighted nodes). Abbreviations: CV_, COVID Visit; DG_, Diagnosis; EP_, Endpoints; IM_, In-house medication; LB_, Lab Results; PC_, Procedures, VS_, Vital Signs; CRP, C-reactive protein.

**Table 2.** Features with significant relationship to decreased mortality. Abbreviations: DG_, Diagnosis; IM_, In-house medication

| Network Name | Feature Name | p-value | OR |
| --- | --- | --- | --- |
| Inpatients post-COVID-19+ PCR Test | DG_>28d_postCOVID \| Z20.828 \| Contact with and (suspected) exposure to other viral communicable diseases | 2.00E-07 | 0.16 |
| Inpatients age < 60 | IM_>28d_postCOVID \| ondansetron | 0.001 | 0.079 |
| Inpatients age < 60 | IM_upto7d_postCOVID \| ondansetron | 0.03 | 0.45 |

### Confirmation of Variables Associated with COVID-19 Mortality

To confirm the association of ondansetron with decreased COVID-19 mortality we ran a logistic regression using 25 variables of interest associated with mortality, including ondansetron use, in the inpatient cohort. The following covariates were significant (Table 3A): *Age* (approximate patient age; p = 5.0e-08), *Ondansetron* (ondansetron use; p = 0.0125), *Heart_Failure* (p = 0.043), *Neoplastic_Disease* (p = 0.0306), *BUN* (p = 1.82e-05) and *onVentilator* (ventilator use; p < 2e-16). These results were obtained by using data from 2,259 patients with complete observations for all covariates used for the analysis.

**Table 3.** Coefficients of logistic regression fitted to original data (panel A) and to five versions of imputed data (panel B). Coefficients with mean p < 0.05 in both original and imputed data sets are bolded. Coefficients with mean p < 0.05 only in one dataset are italicized. Negative sign means association with decreased mortality.

|  | A. Original Data (N=2,259) |  |  | B. Imputed Data (5 sets of N=3,082) |  |  |
| --- | --- | --- | --- | --- | --- | --- |
| Covariate | Estimate | SE | P-value | mean | SD | P-value |
| (Intercept) | -7.690E+00 | 7.41E-01 | < 2E-16 | -7.26E+00 | 4.18E-02 | 1.57E-29 |
| **DM_age_band_label** | **5.06E-02** | **9.28E-03** | **5.00E-08** | **4.70E-02** | **4.18E-04** | **1.23E-08** |
| DM_patient_gender | 3.22E-01 | 1.94E-01 | 9.61E-02 | 2.43E-01 | 5.29E-03 | 1.56E-01 |
| DM_Race_White | -1.228E-01 | 1.861e-01 | 5.09E-01 | -1.66E-01 | 1.60E-02 | 3.20E-01 |
| DM_Ethnicity_Hispanic | 3.74E-02 | 1.92E-01 | 8.46E-01 | 6.82E-02 | 4.65E-02 | 6.98E-01 |
| **Ondansetron** | **-5.44E-01** | **2.180e-01** | **1.25E-02** | **-6.30E-01** | **1.08E-02** | **1.76E-03** |
| Azithromycin | -1.02E-01 | 2.000e-01 | 6.12E-01 | -1.59E-01 | 1.04E-02 | 3.85E-01 |
| *Remdesivir* | *4.11E-01* | *2.11E-01* | *5.17E-02* | *4.00E-01* | *1.32E-02* | *3.47E-02* |
| Dexamethasone | -2.42E-01 | 2.245e-01 | 2.81E-01 | -2.62E-01 | 8.75E-03 | 2.18E-01 |
| Tocilizumab | 1.09E-01 | 2.25E-01 | 6.28E-01 | -3.69E-02 | 1.24E-02 | 8.65E-01 |
| Convalescent_plasma | 1.98E-01 | 2.25E-01 | 3.80E-01 | 1.99E-01 | 6.62E-03 | 3.49E-01 |
| Diabetes | 1.06E-02 | 1.95E-01 | 9.57E-01 | -5.93E-02 | 6.68E-03 | 7.34E-01 |
| COPD | 1.88E-01 | 2.54E-01 | 4.60E-01 | 3.30E-01 | 6.98E-03 | 1.36E-01 |
| Asthma | 7.44E-02 | 3.02E-01 | 8.05E-01 | -4.99E-02 | 1.10E-02 | 8.59E-01 |
| CAD | 2.82E-01 | 2.19E-01 | 1.98E-01 | 2.60E-01 | 4.34E-03 | 1.90E-01 |
| **Heart_Failure** | **4.35E-01** | **2.15E-01** | **4.31E-02** | **4.74E-01** | **1.17E-02** | **1.38E-02** |
| **Neoplastic_Disease** | **-9.06E-01** | **4.190e-01** | **3.06E-02** | **-7.59E-01** | **1.92E-02** | **4.06E-02** |
| Kidney_Disease | -1.41E-01 | 2.237e-01 | 5.29E-01 | -1.97E-02 | 1.08E-02 | 9.22E-01 |
| CRP | 1.02E-03 | 9.17E-04 | 2.65E-01 | 1.10E-03 | 1.25E-04 | 1.92E-01 |
| *D-dimer* | *3.42E-02* | *1.97E-02* | *8.20E-02* | *4.35E-02* | *6.04E-03* | *1.29E-02* |
| ALT | -2.48E-03 | 2.105e-03 | 2.39E-01 | -2.01E-03 | 1.09E-04 | 2.72E-01 |
| *Ferritin* | *4.70E-05* | *2.50E-05* | *6.02E-02* | *4.93E-05* | *4.33E-06* | *2.06E-02* |
| AST | 1.49E-03 | 1.43E-03 | 2.98E-01 | 1.50E-03 | 6.14E-05 | 2.37E-01 |
| **BUN** | **1.87E-02** | **4.37E-03** | **1.82E-05** | **1.76E-02** | **1.14E-04** | **3.35E-06** |
| Lymphocytes | -1.08E-01 | 1.271e-01 | 3.94E-01 | -1.56E-01 | 5.45E-03 | 1.81E-01 |
| **onVentilator** | **2.81E+00** | **2.107e-01** | **< 2E-16** | **2.83E+00** | **1.41E-02** | **5.52E-50** |

Next, the effect of increasing the size of our dataset by imputing data for variables with missing values was examined. This was accomplished using the predictive mean matching method, which selects a small number of complete observations most similar to and most likely representative of missing values, and randomly replaces the missing values. Following this approach, five imputed datasets were generated and input to the logistic regression described previously. Using the five variants of the larger dataset with imputed data yielded greater statistical power in demonstrating association between the above-mentioned variables and death (Table 3B) [e.g., *Age* (p = 1.23E-08), *Ondansetron* (p = 1.76E-03), *Heart_Failure* (p = 1.38E-02), *Neoplastic_Disease* (p = 4.06E-02), *BUN* (p=3.35E-06) and *onVentilator* (p = 5.52E-50)]. With more observations present, additional variables reached significance: *Remdesivir* (p = 3.47E-02), *D-dimer* (p = 1.29E-02), and *Ferritin* (p = 2.06E-02). The coefficients’ sign indicates that increased age, remdesivir use, heart failure, D-dimer, ferritin, BUN, and being on ventilator are each, independently positively associated with mortality, while ondansetron use and neoplastic disease are each negatively associated with mortality. A descriptive statistic of the ondansetron-treated vs. non-treated cohort is presented in Supp. Table 5.

### Ondansetron use in mechanically ventilated patients is associated with decreased mortality after adjusting for interactions

Because the covariates identified by our model are known to interact with each other, efforts were focused on further analysis to fit a more complex model that included all possible interaction terms. In addition to the 25 main terms, 300 pairwise terms and 7 square terms for the continuous terms (lab values and age) were included. The logistic regression was unable to fit this large number of potential variables to the data. This necessitated the use of a data regularization approach. Essentially, the model is penalized against including many covariates and this results in an optimal solution that is unique.

We used the Elastic Net regression method, a combination of the LASSO and ridge regression methods to validate previous findings related to mortality within 30 days of COVID-19 positive PCR test [11]. Optimization of the *alpha* and *lambda* parameters demonstrated that setting *alpha* = 1 minimizes the error of the fit for all *lambda* values. The alpha parameter determines the relative contribution of LASSO and ridge regression penalties to Elastic Net, so setting alpha = 1 simplified Elastic Net regression to LASSO regression. The optimal *lambda* value was selected by a cross-validation approach as provided by the R package **glmnet**. Using five versions of the dataset with imputed data resulted in the generation of five models (Table 4). The covariates *Age, Heart_Failure, D-dimer, Ferritin, BUN* and *onVentilator* were identified by LASSO in all five versions of the imputed dataset; all with positive coefficients, indicative that they associated with increased mortality at 30 days. The covariate *Ondansetron* was selected in two of the five datasets, with negative coefficients, again confirming the potential reduction in mortality at 30 days (i.e., survival benefit). LASSO identified additional covariates that were observed only once out of the five datasets, the interaction term *Ondansetron:onVentilator* (with a negative coefficient) being one of them. These results suggested that the stochastic aspects of the data imputation and model fitting results in minor variability in the composition of the final model.

**Table 4.**
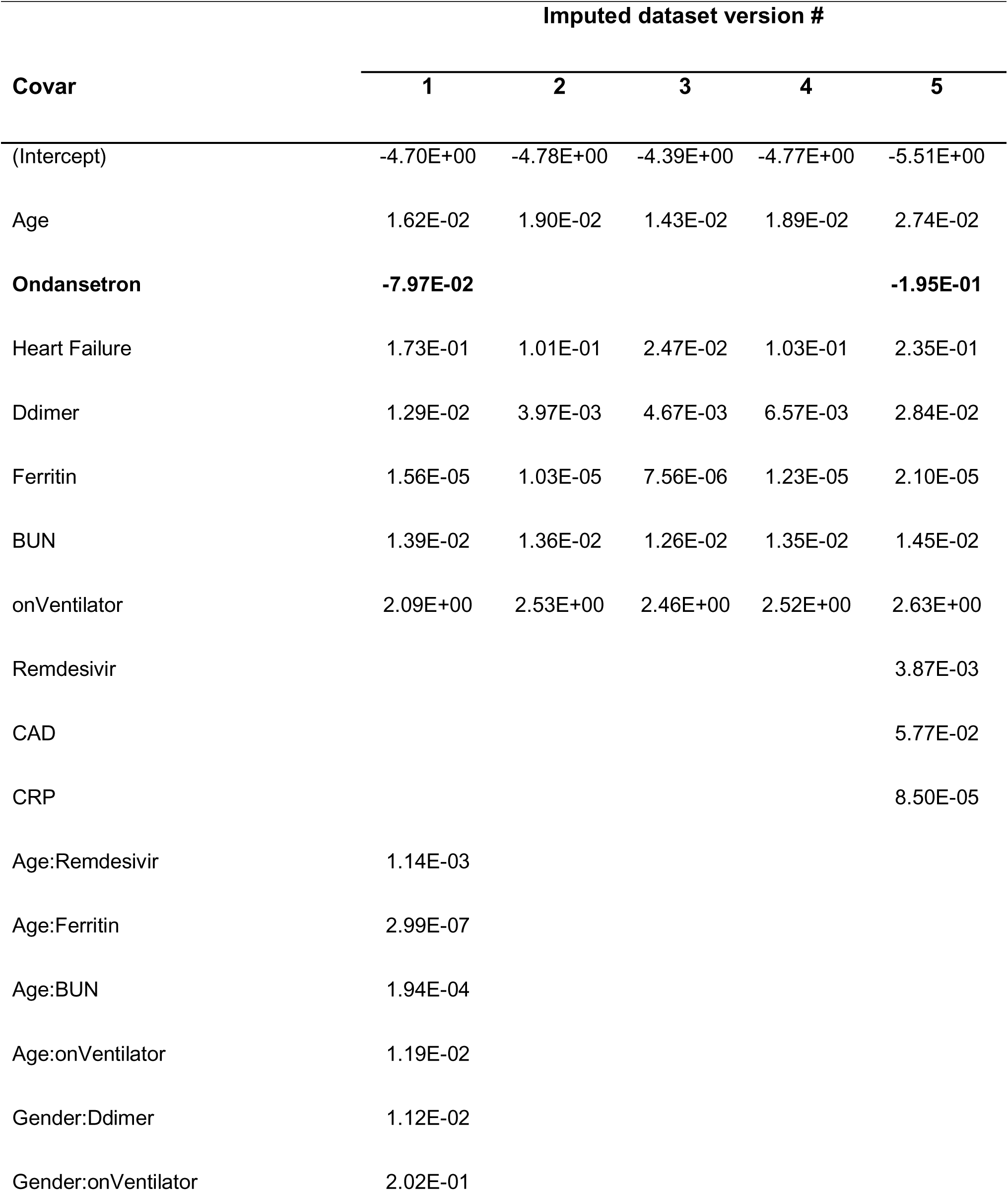
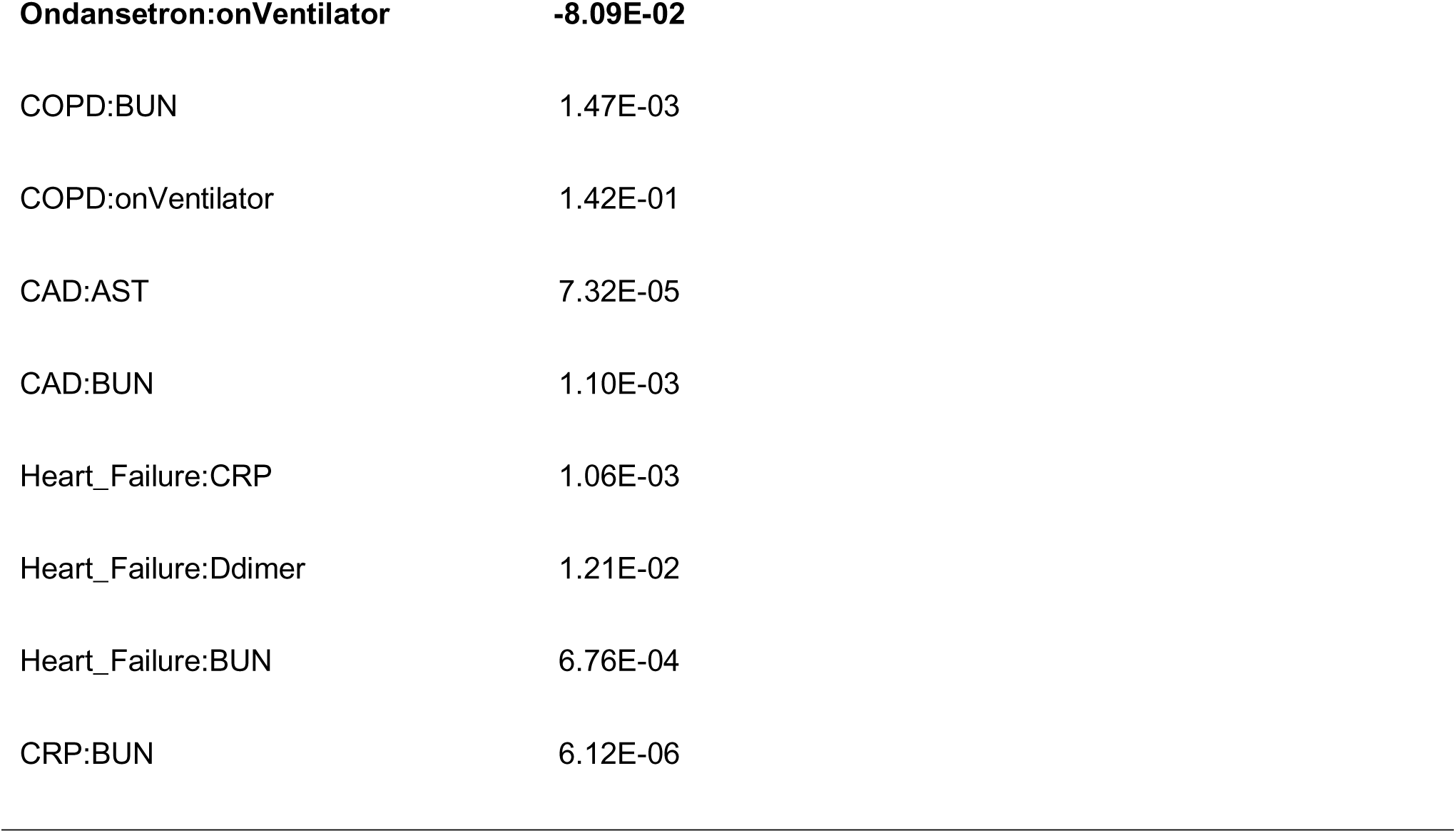
Covariates and their coefficients selected by LASSO (least absolute shrinkage and selection operator) regression on 5 versions of the datasets with imputed data. Negative sign means association with decreased mortality (bold). Abbreviations: Age, approximate age of patient (one of 35, 45, … , 75); AST, aspartate transaminase level; BUN, blood urea nitrogen level; CAD, coronary artery disease; COPD, Chronic obstructive pulmonary disease; CRP, C-reactive protein; (:) indicates interaction between two covariates.

| **Covar** | **Imputed dataset version #** |  |  |  |  |
| --- | --- | --- | --- | --- | --- |
|  | **1** | **2** | **3** | **4** | **5** |
| (Intercept) | -4.70E+00 | -4.78E+00 | -4.39E+00 | -4.77E+00 | -5.51E+00 |
| Age | 1.62E-02 | 1.90E-02 | 1.43E-02 | 1.89E-02 | 2.74E-02 |
| **Ondansetron** | **-7.97E-02** |  |  |  | **-1.95E-01** |
| Heart Failure | 1.73E-01 | 1.01E-01 | 2.47E-02 | 1.03E-01 | 2.35E-01 |
| Ddimer | 1.29E-02 | 3.97E-03 | 4.67E-03 | 6.57E-03 | 2.84E-02 |
| Ferritin | 1.56E-05 | 1.03E-05 | 7.56E-06 | 1.23E-05 | 2.10E-05 |
| BUN | 1.39E-02 | 1.36E-02 | 1.26E-02 | 1.35E-02 | 1.45E-02 |
| onVentilator | 2.09E+00 | 2.53E+00 | 2.46E+00 | 2.52E+00 | 2.63E+00 |
| Remdesivir |  |  |  |  | 3.87E-03 |
| CAD |  |  |  |  | 5.77E-02 |
| CRP |  |  |  |  | 8.50E-05 |
| Age:Remdesivir | 1.14E-03 |  |  |  |  |
| Age:Ferritin | 2.99E-07 |  |  |  |  |
| Age:BUN | 1.94E-04 |  |  |  |  |
| Age:onVentilator | 1.19E-02 |  |  |  |  |
| Gender:Ddimer | 1.12E-02 |  |  |  |  |
| Gender:onVentilator | 2.02E-01 |  |  |  |  |

|  |  |
| --- | --- |
| **Ondansetron:onVentilator** | **-8.09E-02** |
| COPD:BUN | 1.47E-03 |
| COPD:onVentilator | 1.42E-01 |
| CAD:AST | 7.32E-05 |
| CAD:BUN | 1.10E-03 |
| Heart_Failure:CRP | 1.06E-03 |
| Heart_Failure:Ddimer | 1.21E-02 |
| Heart_Failure:BUN | 6.76E-04 |
| CRP:BUN | 6.12E-06 |

To delineate further the basis of the observed variability of the LASSO regression model, and to generate confidence intervals for the generated coefficients, 10 versions of the dataset with imputed values were generated, and each of them in turn was used to generate a population of 1,000 datasets with similar underlying distributions, by the bootstrap method. We ensured that each bootstrap sample was balanced in terms of proportion of patient mortality. The 10,000 samples were each analyzed by LASSO regression. The results are summarized in Table 5, which lists the percentage of the 10,000 bootstrap samples for the top covariates selected by LASSO, their median coefficient values, as well as their 95% and 99% confidence intervals.

**Table 5.** Covariates selected by logistic LASSO (least absolute shrinkage and selection operator) regressions in greater than 50% of 1000 bootstrap permutations of 10 versions of the imputed dataset. Covariates are sorted by the median value of their coefficients, along with their 95% and 99% confidence intervals (CI). Negative median sign means association with decreased mortality (bold). Age, (approximate age (one of 35, 45, …, 75); Abbreviations: BUN, blood urea nitrogen level; COPD, Chronic obstructive pulmonary disease; CRP, C-reactive protein; (:) indicates interaction between two covariates.

| Name | Percent<br>selected | Coefficient |  |  |
| --- | --- | --- | --- | --- |
|  |  | Median | CI 99% | CI 95% |
| (Intercept) | 100 | -5.10E+00 | (-6.03, -3.92) | (-6.31, -3.57) |
| **Tocilizumab:onVentilator** | **56.3** | **-5.00E-01** | **(-1.59, -0.0213)** | **(-1.94, -4.06e-03)** |
| **Gender:Neoplastic_Disease** | **61.7** | **-4.87E-01** | **(-1.69, -0.0274)** | **(-2.1, -4.98e-03)** |
| **Ondansetron:onVentilator** | **74.4** | **-3.65E-01** | **(-1.14, -0.0201)** | **(-1.38, -4.51e-03)** |
| Age:Ferritin | 55.3 | 3.47E-07 | (2.60e-09, 1.22e-06) | (2.06e-08, 9.57e-07) |
| CRP:BUN | 68 | 2.80E-05 | (2.03e-07, 1.17e-04) | (1.42e-06, 9.15e-05) |
| Age^2 | 99.1 | 2.68E-04 | (3.25e-05, 5.00e-04) | (7.95e-05, 4.42e-04) |
| DM_age_band_label:Remdesivir | 75.9 | 3.67E-03 | (3.75e-05, 1.68e-02) | (2.36e-04, 1.29e-02) |
| BUN | 52.9 | 7.39E-03 | (2.95e-05, 2.23e-02) | (2.27e-04, 1.84e-02) |
| Gender:Ddimer | 60.6 | 2.37E-02 | (1.70e-04, 8.38e-02) | (1.09e-03, 6.79e-02) |
| Gender:onVentilator | 61.4 | 2.30E-01 | (1.34e-03, 9.10e-01) | (9.56e-03, 7.19e-01) |
| COPD:onVentilator | 51.3 | 2.64E-01 | (-1.36e-01, 1.25e+00) | (7.95e-03, 9.44e-01) |
| Ondansetron:Convalescent_plasma | 52.9 | 4.56E-01 | (2.94e-03, 1.69e+00) | (2.33e-02, 1.34e+00) |
| onVentilator | 99.8 | 2.54E+00 | (4.73e-01, 3.76e+00) | (1.07e+00, 3.41e+00) |

The LASSO regression results on the bootstrap samples show *BUN* and *onVentilator* identified as main (linear) terms, while the remaining covariates are interaction terms. *Age* is a quadratic (squared) term, indicating that mortality increases curvilinearly with age. The most frequently identified covariates are *onVentilator* and *Age^2* and both are positively associated with mortality at 30 days. The interaction term *Ondansetron:onVentilator* is identified in 74.4% of the bootstrap sample and is negatively associated with mortality. None of the 95% confidence intervals, and except for *COPD:onVentilator*, none of the 99% confidence intervals of the coefficients include zero, suggesting that these coefficients are stable in their sign despite the variability in the sample sets.

From the regression analysis we conducted on the bootstrapped samples of the dataset, the median value of the regression coefficient for mechanically ventilated patients treated with ondansetron was -0.365. This means that when this interaction term has a value of 1, the odds of death are multiplied by e^(-0.365)^ = 0.694.

## DISCUSSION

The goal of this study, representing a multi-institutional collaborative effort of collecting, structuring, and analyzing RWD through AI analytics, was to generate insights on COVID-19 mortality-associated factors and identify potential new therapeutic options for COVID-19 patients. The study involved two stage data analytics: the primary discovery phase, involving Bayesian statistics-based analysis to generate causal networks of all patients to identify significant factors influencing mortality in the COVID-19 positive cohort (Table 1), from 12 months pre- to 28 days post PCR-based COVID-19 diagnosis; this was followed by confirmatory tests of the main findings on imputed and bootstrapped data, from multivariable regression analysis to LASSO logistic regression. The Bayesian network findings were based on the analysis of demographic, clinical, and laboratory data from 16,277 PCR confirmed COVID-19 patients representing a subset of the 279,281 patients in the RECOVER-19 registry; the confirmatory logistic regression analyses were performed on data from the 3,082 hospitalized patients.

We identified ondansetron as the main factor associated with improved survival in mechanically ventilated COVID-19 patients. An initial unbiased search for predictors of mortality at any time and within any patient population found ondansetron as the only medication associated with decreased mortality (Table 2). This association was initially identified within a specific inpatient population (< 60 years old) and when ondansetron was administered at disparate times (up to 7 days and > 28 days post COVID-19 diagnosis). Regarding the timing of administration, of the 737 patients who received ondansetron at any time during the first 30 days, the majority (84%) received it within the first week post SARS-CoV-2 positive PCR test. Patients who received ondansetron in the first week post PCR test had improved survival compared with patients who did not (p < 0.0001) (Figure 4). Multivariable logistic regression by LASSO showed significant effects for age and ondansetron use on mortality within 30 days, but not an *Age:Ondansetron* interaction effect, meaning that the ondansetron effect applies to all age groups equally. However, it is the interaction term *Ondansetron:onVentilator* that is primarily selected by LASSO as a covariate of non-zero coefficient, rather than the main term *Ondansetron*. This indicates that the beneficial effect of ondansetron is seen only in patients on ventilator. This key finding complements a study by Bayat et. al. that reported a reduction in 30-day all-cause mortality for all inpatients (including ICU) with early administration of ondansetron after admission [12].

**Figure 4.**
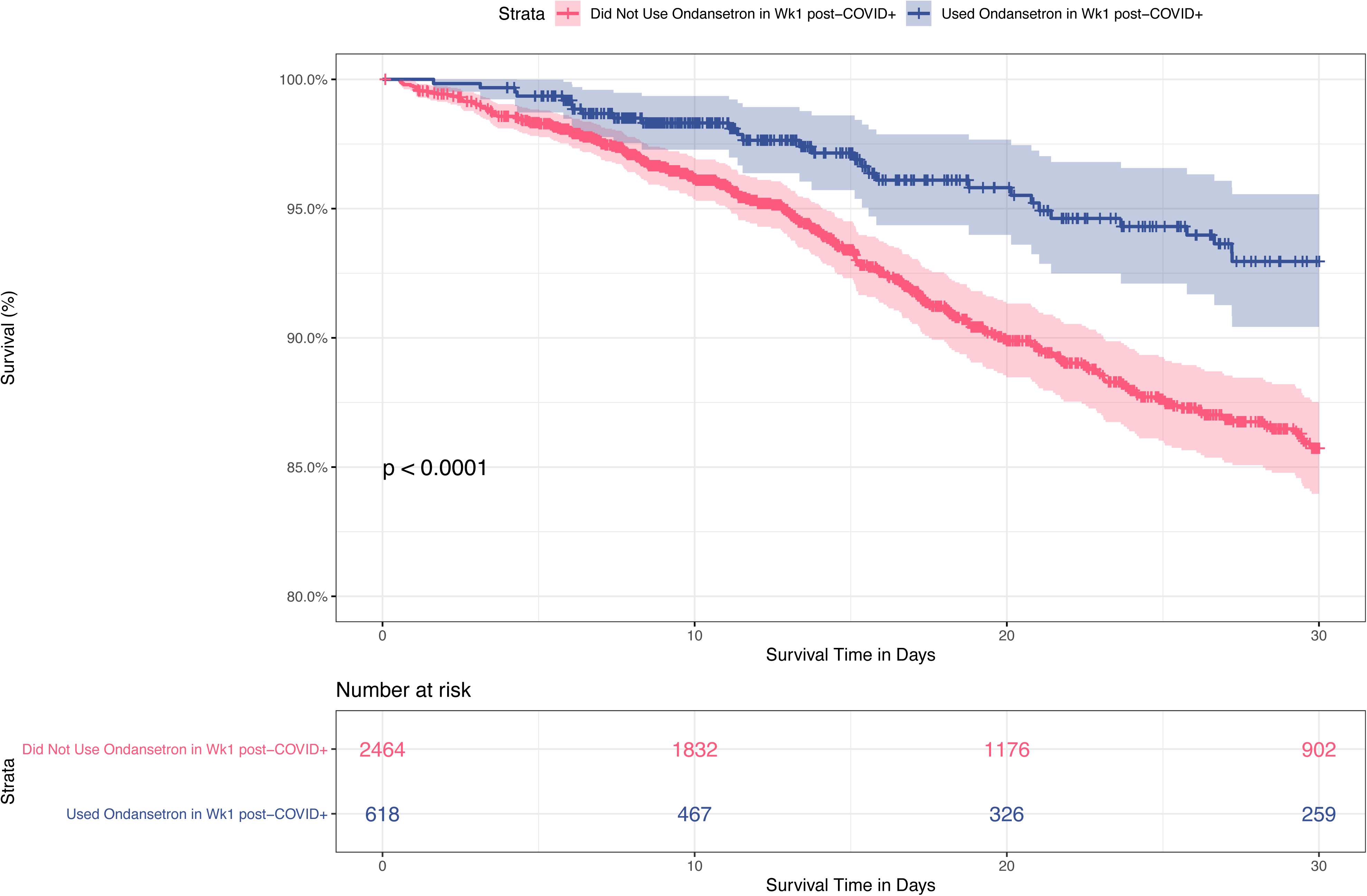
Kaplan-Meier curve showing 30-day survival rates of hospitalized patients who received (blue) or did not receive (red) ondansetron in the first week post SARS-CoV-2 positive PCR test. Patients who received ondansetron had improved 30-day survival compared with patients who did not (p < 0.0001).

Ondansetron is a selective 5-HT3 serotonin-receptor antagonist with known effects against nausea and vomiting through both central and peripheral mechanisms [13]. It has been postulated that SARS-CoV-2 might have an indirect effect on enteroendocrine cells (EEC), triggering the release of neuroactive agents such as the emesis inducing serotonin [14]. Most studies showed that patients with COVID-19 have higher plasma serotonin levels and this correlates with increased IL-6 [15, 16], while others concluded they have decreased serotonin levels [17]. Considering serotonin’s role in regulating innate and adaptive immune responses [18], the observed beneficial effect of ondansetron might be due to the modulation of serotonin levels or could also be linked with a direct effect on the immune system [19] or on known COVID-19 comorbidities, such liver and kidney disease or complications, such as thrombosis [20–22]. There are also data suggesting that serotonin receptor signaling influences cellular activities that regulate entry of diverse virus families [23].

It is interesting to note that while we do not find convalescent plasma to be a significant predictor of death, patients on ondansetron and convalescent plasma were more likely to die (Table 5), suggesting a complex interaction between ondansetron, ventilator use, and convalescent plasma. It is now known that high-titer convalescent plasma does not improve COVID-19 survival or clinical outcomes when used in both inpatients [24, 25] and high-risk outpatients [26] and when a beneficial effect on the risk of death was observed it was not maintained for patients who had received mechanical ventilation [27]. As, early in the pandemic, convalescent plasma was usually reserved for patients with more severe COVID-19 pneumonia, this observed association might be explained by this confounding bias and not by a potential detrimental effect of convalescent plasma.

In our cohort, tocilizumab had a similar effect on mortality as ondansetron for mechanically ventilated patients, in line with published evidence from large, randomized control studies [28, 29].

Although being male is associated with COVID-19 mortality, we find that in the context of neoplastic disease this is reversed. Our results show a negative association between the interaction term *Gender:Neoplastic_Disease* and mortality. This may be due to an indirect ondansetron effect, since this is often prescribed to cancer patients undergoing chemotherapy, radiation therapy, and surgery. So, while being male is positively associated with COVID-19 mortality, in cancer patients this may be modulated by ondansetron use. Also, the association of COVID-19 mortality with cancer is not straightforward. The COVID-19 mortality of cancer patients depends on the type of their cancer, with the main mortality drivers being age, gender, comorbidities, and hematological cancers [30–32].

In the present study, besides ondansetron and tocilizumab, other covariates interacting with ventilator use indicate that males on ventilator and patients with chronic obstructive pulmonary disease (COPD) on ventilator are more likely to die. The former of these two findings agrees with Nicholson et al., who showed that male COVID-19 patients on ventilator have a higher mortality rate than females (after correcting for co-morbidities) [33]. COPD is also an already established comorbidity associated with increased odds of hospitalization and death in COVID-19 patients [34, 35].

Looking at interactions between other covariates, we found that the CRP and BUN dyad, laboratory biomarkers that are found in prognostic models for COVID-19 mortality, are also associated with mortality in our cohort [36, 37]. Similarly, the previously observed mortality link of interacting factors ferritin and age [38] as well as higher D-dimer levels in males were also confirmed by our analyses [39]. The association between mortality with a combination of age and age-squared agrees with a previous finding that infection fatality ratio has a log-linear increase by age among individuals older than 30 years [40].

Although an FDA-approved drug for COVID-19, remdesivir was not found to increase survival in large, randomized control trials [41–43]. We find that age and remdesivir use interact to increase mortality. This has not been reported previously and may suggest that we see a similar confounding bias with convalescent plasma, since remdesivir was reserved for more severe patients earlier on.

Diagnostic code Z20.828 (“Contact with and (suspected) exposure to other viral communicable diseases”) was one of the 3 features with a significant relationship with decreased mortality in our analysis. This code was used in 2020 when a clinician suspected exposure to SARS-CoV-2 without a test result available. In the RECOVER-19 registry, out of the approximately 200,000 unique patients seen with this diagnostic code in 2020, about 16,000 were found to be positive. We might see this mortality benefit because the SARS-CoV-2 positive patients with the Z20.828 code might have had a less severe form of COVID-19 (higher ambiguity without a positive test) or arrived earlier in the course of the disease and were designated patients under investigation (PUI), benefiting from early precautions. Generally, the patients admitted with a severe form of COVID-19 would have received another more definitive diagnostic code.

## CONCLUSION

To our knowledge, we are the first to use Bayesian network analysis of clinical data to report disease outcomes in COVID-19 patients. Using high-performance computing driven Bayesian AI, we report here a negative association between mortality and ondansetron treatment for mechanically ventilated patients, as well as confirming the beneficial effects of tocilizumab and validating some of the already established factors associated with COVID-19 increased mortality, such as higher BUN, CRP, ferritin, and D-dimer levels. These results confirm the validity of our approach and the hypothesis-generating potential of the bAIcis® platform. Currently there are no controlled trials examining the effect of ondansetron in COVID-19 patients. Our findings suggest that this FDA approved drug should be investigated for its potential effectiveness against COVID-19.

## Data Availability

De-identified data is available per request to the corresponding author

## SUPPLEMENTAL METHODS

### Data Sources

The AdventHealth big data platform is a Dedicated Cloudera Distribution Big Data cluster housed on site in Oracle Big Data Appliance (BDA). This BDA houses the IDEA platform, which has a Data Lake and an Enterprise Data Warehouse. The Data Lake is catalogued by source system and additional subject area specific conformed repositories are interconnected with Enterprise level dimensional data. This architecture contains near real time data (4 to 5 second latency) feeds from over 1,000 source data tables. A Kafka footprint is also in place to support near real time data streaming. Based on the various source systems the feed timing can be near real time, daily, weekly, monthly, quarterly or on demand. There is a wide variety of data sources feeding this repository but some of the larger footprints are near real time CERNER EHR data, Athena EHR Data Warehouse feed Version 17.1, Population Health Claims data, and Patient Experience. The Data Lake is not only capable of ingesting traditional data, but it also supports semi-structured and unstructured data, including FHIR data.

The COVID-19 data analytics effort is supported by the AdventHealth IDEA scalable distributed data platform. Briefly, raw data from the EHR systems are ingested into the Data Lake. No modeling is done at this stage, but only minimal data processing such as merging EHR data from all geographical domains and conforming date/time elements to a common local time-zone. This allows multiple use cases for research, providing an almost real-time understanding of the size and distribution of the COVID-19 patient cohort across all 53 AdventHealth hospitals. A shared curated data set specific to COVID-19 was created to centralize the collection effort of key subject areas to include patient, encounters, and clinical data.

The Clinical Classifications Software Refined (CCSR) is a publicly available taxonomy created and maintained by the Agency for Healthcare Research and Quality which aggregates International Classification of Diseases, 10th Revision, Clinical Modification/Procedure Coding System (ICD-10-CM/PCS) codes into clinically meaningful categories.

A COVID-19 positive PCR test was defined as a lab result in which (1) *PCR_order_flag* = 1, (2) *result_positive_flag* = 1, and (3) *nomenclature_short_string* was explanatory for SARS-CoV-2 testing.

Patients were defined as deceased if having *deceased_flag* = 1 in the patients table, or if having had an encounter with *discharge_disposition* = “Expired – 20.” Being placed on a ventilator was defined as having a procedure with *ccs_catg_dsc* = “Respiratory intubation and mechanical ventilation” that occurred at the time of PCR sample specimen collection or later. Similarly, being admitted to Inpatient care was defined as having an encounter with *enctr_type_class* = “Inpatient” at the time of PCR sample specimen collection or later. A patient was defined as having been admitted to the ICU if a patient had a lab test occurring at the time of PCR sample collection or later with (1) *nomenclature_short_string* = “Admitted to ICU for condition” and (2) *result_value* = “YES.”

LOS was defined using the following criteria: First, a COVID-19 positive encounters was defined as an encounter in which *enc_c19pos* = 1. Next, these COVID-19 positive encounters were considered valid for LOS calculation if the *discharge_dttm* was specified. If a patient had one or more valid COVID-19 positive encounters, LOS was taken to be the sum of LOS values for each valid COVID-19 positive encounter. By contrast, patients who had only invalid COVID-19 positive encounters (i.e., COVID-19 positive encounters are listed, but all encounters have unspecified *discharge_dttm*) had LOS defined as not available. Finally, any patient who had no COVID-19 positive encounters (valid or invalid) had LOS defined as zero.

### Feature derivation

Diagnosis features within each time bin were defined as the presence or absence of diagnoses extracted from the diagnosis table. Any patient with a listed ICD10 code (*icd10_cm_cd*) at a time (*active_status_dt_tm*) within a defined time bin was set to TRUE for this diagnosis feature in this time window, whereas this corresponding diagnosis feature was set to FALSE for all other patients in this time window. Infrequently occurring diagnosis features were aggregated using the **icd** library using R package version 4.0.9. (https://jackwasey.github.io/icd/). Patients who were discharged before the beginning of a particular time window had all diagnosis features for this time window assigned as missing.

Similarly, problem features within each time bin were defined as the presence or absence of problems extracted from the Problems table. First, only active entries in the problems table (*life_cycle_status* = Active) were used for feature extraction. Next, any patient with a listed SNOMED code (s*nomed_ct_code*) at a time (b*eg_effective_dt_tm*) within a defined time bin was set to TRUE for this problem feature, while this problem feature was set to FALSE for all other patients in this time window. Finally, patients who were discharged before the beginning of a particular time window had all problem features for this time window assigned as missing.

In-house medication features and home medication features for each time bin were derived from the InHouse Medications and Home Medications tables, respectively. For in-house medication features, a value of TRUE was assigned if a patient received an in-house medication (*med_name*) at a time (*start_date_time*) within a specified time bin; otherwise, a value of FALSE was assigned. Notably, in-house medications with name “premix diluent” were excluded from in-house medication feature generation. Similarly, for home medication features, a value of TRUE was assigned if a patient was prescribed a medication (*ordered_drug*) at a time (*req_start_dttm*) within a specified time bin, else a value of FALSE was assigned for patients who didn’t receive this medication within this time bin. Patients who were discharged before the beginning of a particular time window had all in-house medication and home medication features for this time window assigned as missing.

Procedure features for each time bin were derived from the Procedures table in a similar approach to the diagnosis features. For procedure features, a value of TRUE was assigned if a patient underwent a procedure (icd10_pcs_cd) within a time window of interest (*active_status_dt_tm*), while patients who did not were assigned a value of FALSE. Infrequent procedure features were aggregated using the **icd** library. Patients who were discharged before the beginning of a particular time window had all procedure features for this time window assigned as missing.

Lab value features for each time bin were derived from the Lab Results table. First, any entries in the Lab Results table with undefined numeric values (*result_value_numeric*) or normalcy listed as “No Flag” were excluded. Next, exploratory data analysis found that lab results with unspecified normalcy were in the normal range, thus lab results with unspecified normalcy were considered normal. Lab value features were then defined corresponding to each lab test LOINC code (*loinc*) with description (*nomenclature_short_string*) measured for a given patient within a time window of interest (*lab_order_date*), with features values taken to be the lab value normalcy (Normal, Abnormal, High, Low, or Critical). If a particular lab value was measured more than once for a given patient in a time window of interest, the lab values measured on the nearest date to the date of COVID-19+ PCR specimen collection were used. If multiple tests were reported on this date, the lab result feature was defined as the most severe lab result measured, according to a ranking of Critical > High > Low. Finally, patients who did not have recorded lab values in this time window had their lab value features assigned as missing.

In a similar manner, vital status features for each time bins were derived from the Clinical Events table. First, any entries in the Clinical Events table with normalcy set to “No Flag” were excluded. Additionally, entries with value zero (*result_val_numeric*) and unspecified normalcy were excluded. As had been shown with Lab values, exploratory data analysis found that entries with unspecified normalcy were within the normal range, therefore entries with unspecified normalcy were considered normal. Vital status features were then defined corresponding to each event code (*event_cd*) and event description (*event_desc*) measured for a given patient within a time window of interest (*valid_from_dt_tm*), with feature values taken to be the vital sign normalcy (Normal, Abnormal, High, or Low). To reduce the number of feature levels, entries with normalcy outside the test bounds (<LLOW and >HHI) were considered LOW and HI, respectively. Vital sign measurements that were listed more than once for a given patient in a time window of interest, were summarized by using the vital sign measured on the nearest date to the date of COVID-19 positive PCR specimen collection. If multiple vital signs were reported on this date, the vital sign feature was defined as the most severe vital sign result measured, according to a ranking of HI > LOW > ABN. Finally, as with the derivation of lab test features, patients who did not have recorded vital sign values in this time window had their vital sign features assigned as missing.

### Elastic Net regression method

Optimization of the *alpha* and *lambda* parameters demonstrated that setting *alpha* = 1 minimizes the error of the fit for all *lambda* values. The *alpha* parameter determines the relative contribution of LASSO and ridge regression penalties to Elastic Net, so setting *alpha* = 1 simplified Elastic Net regression to LASSO regression. The optimal *lambda* value was selected by a cross-validation approach as provided by the R package **glmnet**.

### Bootstrapping

To delineate further the basis of the observed variability of the LASSO regression model, and to generate confidence intervals for the generated coefficients, 10 versions of the dataset with imputed values was generated, and each of them in turn was used to generate a population of 1000 datasets with similar underlying distributions, by the bootstrap method. We ensured that each bootstrap sample was balanced in terms of the number of patient-specific mortality. The 10,000 samples were each analyzed by LASSO regression.

## SUPPLEMENTAL TABLE LEGEND

**Supplemental Table 1.** Registry and Biorepository of COVID-19 for AdventHealth (RECOVER-19) data tables. Abbreviations: SARS-CoV-2, severe acute respiratory syndrome coronavirus 2; COVID-19, coronavirus disease 2019; ICD-10-CM, International Classification of Diseases, Tenth Revision, Clinical Modification; SNOMED-CT, Systematized Nomenclature of Medicine Clinical Terms; PCS, Procedure; LOINC, Logical Observation Identifiers Names and Codes.

**Supplemental Table 2.** Features with significant relationship to increased mortality. Abbreviations: CV_, COVID Visit; DG_, Diagnosis; EP_, Endpoints; IM_, In-house medication; LB_, Lab Results; PC_, Procedures, VS_, Vital Signs

**Supplemental Table 3.** Significant predictors of mortality in 19 networks generated by BERG’s Bayesian AI analytics (bAIcis®) learning

**Supplemental Table 4.** Relationship between LOS and mortality. Note that p-values were computed using t-test. EP_, endpoints.

**Supplemental Table 5.** Descriptive statistic of ondansetron treated vs. non treated patients.

## Acknowledgements

We would like to thank Lance McCain for his leadership and support in establishing the AdventHealth RECOVER-19 registry

## REFERENCES

1. Johns Hopkins University (JHU). COVID-19 Dashboard by the Center for Systems Science and Engineering (CSSE) at Johns Hopkins University (JHU). https://coronavirus.jhu.edu/map.html 2021 [cited 2021 July 21, 2021]; COVID-19 Dashboard by the Center for Systems Science and Engineering (CSSE) at Johns Hopkins University (JHU)].

2. DeGrave, A.J., J.D. Janizek, and S.I. Lee, AI for radiographic COVID-19 detection selects shortcuts over signal. medRxiv, 2020.

3. Xu, W., et al., Risk factors analysis of COVID-19 patients with ARDS and prediction based on machine learning. Sci Rep, 2021. 11(1): p. 2933.

4. Tariq, A., et al., Patient-specific COVID-19 resource utilization prediction using fusion AI model. NPJ Digit Med, 2021. 4(1): p. 94.

5. The Lancet Digital, H., Artificial intelligence for COVID-19: saviour or saboteur? Lancet Digit Health, 2021. 3(1): p. e1.

6. Yu, J., et al., Advances to Bayesian network inference for generating causal networks from observational biological data. Bioinformatics, 2004. 20(18): p. 3594–603.

7. Zhang, L., et al., bAIcis: A Novel Bayesian Network Structural Learning Algorithm and Its Comprehensive Performance Evaluation Against Open-Source Software. J Comput Biol, 2020. 27(5): p. 698–708.

8. Healthcare Cost and Utilization Project. Clinical Classifications Software Refined (CCSR). https://www.hcup-us.ahrq.gov/toolssoftware/ccsr/ccs_refined.jsp 2021 [cited 2021.

9. Rubin, D.B., Statistical Matching Using File Concatenation With Adjusted Weights and Multiple Imputations. Journal of Business & Economic Statistics, 1986. 4(1): p. 87–94.

10. van Buuren, S. and K. Groothuis-Oudshoorn, mice: Multivariate Imputation by Chained Equations in R. 2011, 2011. 45(3): p. 67.

11. Zou, H. and T. Hastie, Regularization and Variable Selection via the Elastic Net. Journal of the Royal Statistical Society. Series B (Statistical Methodology), 2005. 67(2): p. 301–320.

12. Bayat, V., et al., Reduced Mortality With Ondansetron Use in SARS-CoV-2-Infected Inpatients. Open Forum Infect Dis, 2021. 8(7): p. ofab336.

13. Griddine, A. and J.S. Bush, Ondansetron, in StatPearls. 2021: Treasure Island (FL).

14. Andrews, P.L.R., et al., COVID-19, nausea, and vomiting. J Gastroenterol Hepatol, 2021. 36(3): p. 646–656.

15. Ha, S., et al., Serotonin is elevated in COVID-19-associated diarrhoea. Gut, 2021. 70(10): p. 2015–2017.

16. Zaid, Y., et al., Platelets Can Associate with SARS-Cov-2 RNA and Are Hyperactivated in COVID-19. Circ Res, 2020.

17. Soria-Castro, R., et al., SEVERE COVID-19 IS MARKED BY DYSREGULATED SERUM LEVELS OF CARBOXYPEPTIDASE A3 AND SEROTONIN. medRxiv, 2021: p. 2021.02.02.21251020.

18. Hamed, M.G.M. and R.S. Hagag, The possible immunoregulatory and anti-inflammatory effects of selective serotonin reuptake inhibitors in coronavirus disease patients. Med Hypotheses, 2020. 144: p. 110140.

19. Magrini, E., et al., Serotonin-mediated tuning of human helper T cell responsiveness to the chemokine CXCL12. PLoS One, 2011. 6(8): p. e22482.

20. Chen, Y., et al., Association of 5-Hydroxytryptamine 3 Receptor Antagonists With the Prognosis of Liver Failure. Front Pharmacol, 2021. 12: p. 648736.

21. Liu, F.C., et al., The anti-aggregation effects of ondansetron on platelets involve IP3 signaling and MAP kinase pathway, but not 5-HT3-dependent pathway. Thromb Res, 2012. 130(3): p. e84–94.

22. Datta, A., et al., ’Black Box’ to ’Conversational’ Machine Learning: Ondansetron Reduces Risk of Hospital-Acquired Venous Thromboembolism. IEEE J Biomed Health Inform, 2021. 25(6): p. 2204–2214.

23. Mainou, B.A., et al., Serotonin Receptor Agonist 5-Nonyloxytryptamine Alters the Kinetics of Reovirus Cell Entry. J Virol, 2015. 89(17): p. 8701–12.

24. Group, R.C., Convalescent plasma in patients admitted to hospital with COVID-19 (RECOVERY): a randomised controlled, open-label, platform trial. Lancet, 2021. 397(10289): p. 2049–2059.

25. Group, T.C.-S., et al., Convalescent plasma for hospitalized patients with COVID-19 and the effect of plasma antibodies: a randomized controlled, open-label trial. medRxiv, 2021: p. 2021.06.29.21259427.

26. Korley, F.K., et al., Early Convalescent Plasma for High-Risk Outpatients with Covid-19. N Engl J Med, 2021.

27. Joyner, M.J., et al., Convalescent Plasma Antibody Levels and the Risk of Death from Covid-19. N Engl J Med, 2021. 384(11): p. 1015–1027.

28. Group, R.C., Tocilizumab in patients admitted to hospital with COVID-19 (RECOVERY): a randomised, controlled, open-label, platform trial. Lancet, 2021. 397(10285): p. 1637–1645.

29. Mariette, X., et al., Effectiveness of Tocilizumab in Patients Hospitalized With COVID-19: A Follow-up of the CORIMUNO-TOCI-1 Randomized Clinical Trial. JAMA Intern Med, 2021.

30. Lee, L.Y.W., et al., COVID-19 prevalence and mortality in patients with cancer and the effect of primary tumour subtype and patient demographics: a prospective cohort study. Lancet Oncol, 2020. 21(10): p. 1309–1316.

31. Lee, L.Y., et al., COVID-19 mortality in patients with cancer on chemotherapy or other anticancer treatments: a prospective cohort study. Lancet, 2020. 395(10241): p. 1919–1926.

32. Kuderer, N.M., et al., Clinical impact of COVID-19 on patients with cancer (CCC19): a cohort study. Lancet, 2020. 395(10241): p. 1907–1918.

33. Nicholson, C.J., et al., Estimating Risk of Mechanical Ventilation and Mortality Among Adult COVID-19 patients Admitted to Mass General Brigham: The VICE and DICE Scores. medRxiv, 2020.

34. Lee, S.C., et al., Impact of COPD on COVID-19 prognosis: A nationwide population-based study in South Korea. Sci Rep, 2021. 11(1): p. 3735.

35. Gerayeli, F.V., et al., COPD and the risk of poor outcomes in COVID-19: A systematic review and meta-analysis. EClinicalMedicine, 2021. 33: p. 100789.

36. Altschul, D.J., et al., A novel severity score to predict inpatient mortality in COVID-19 patients. Sci Rep, 2020. 10(1): p. 16726.

37. Booth, A.L., E. Abels, and P. McCaffrey, Development of a prognostic model for mortality in COVID-19 infection using machine learning. Mod Pathol, 2021. 34(3): p. 522–531.

38. Rasyid, H., et al., Impact of age to ferritin and neutrophil-lymphocyte ratio as biomarkers for intensive care requirement and mortality risk in COVID-19 patients in Makassar, Indonesia. Physiol Rep, 2021. 9(10): p. e14876.

39. He, X., et al., The poor prognosis and influencing factors of high D-dimer levels for COVID-19 patients. Sci Rep, 2021. 11(1): p. 1830.

40. O’Driscoll, M., et al., Age-specific mortality and immunity patterns of SARS-CoV-2. Nature, 2021. 590(7844): p. 140–145.

41. Beigel, J.H., et al., Remdesivir for the Treatment of Covid-19 - Final Report. N Engl J Med, 2020. 383(19): p. 1813–1826.

42. Consortium, W.H.O.S.T., et al., Repurposed Antiviral Drugs for Covid-19 - Interim WHO Solidarity Trial Results. N Engl J Med, 2021. 384(6): p. 497–511.

43. Wang, Y., et al., Remdesivir in adults with severe COVID-19: a randomised, double-blind, placebo-controlled, multicentre trial. Lancet, 2020. 395(10236): p. 1569–1578.

