## Supplementary File Table 3 for "Ondansetron use is associated with lower COVID-19 mortality in a Real-World Data network-based analysis"

**Supplemental Table 3**

| Feature Name | Number of Networks | Directionality | All Patients | Inpatients | Inpatients pre-COVID-19+ PCR Test | Inpatients post-COVID-19+ PCR Test | Inpatients age 18 to 39 | Inpatients age 40 to 49 | Inpatients age 50 to 59 | Inpatients age 60 to 69 | Inpatients age 70+ | Inpatients age < 60 | Inpatients age 60+ | Inpatients age < 60 pre-COVID-19+ PCR Test | Inpatients age 60+ pre-COVID-19+ PCR Test | Inpatients African American Non-Hispanic | Inpatients Hispanic | Inpatients White Non-Hispanic | Inpatients African American Non-Hispanic pre-COVID-19+ PCR Test | Inpatients Hispanic pre-COVID-19+ PCR Test | Inpatients White Non-Hispanic pre-COVID-19+ PCR Test |
| --- | --- | --- | --- | --- | --- | --- | --- | --- | --- | --- | --- | --- | --- | --- | --- | --- | --- | --- | --- | --- | --- |
| IM_>28d_postCOVID \| ondansetron | 1 | Decreased |  |  |  |  |  |  |  |  |  | X |  |  |  |  |  |  |  |  |  |
| IM_upto7d_postCOVID \| ondansetron | 1 | Decreased |  |  |  |  |  |  |  |  |  | X |  |  |  |  |  |  |  |  |  |
| DG_>28d_postCOVID \| Z20.828 \| Contact with and (suspected) exposure to other viral communicable diseases | 1 | Decreased |  |  |  | X |  |  |  |  |  |  |  |  |  |  |  |  |  |  |  |
| EP_placedOnVentilator | 16 | Increased | X | X | X | X |  | X |  | X | X | X | X | X | X |  | X | X | X | X | X |
| EP_admittedToICU | 11 | Increased | X |  | X |  |  |  |  | X | X |  | X | X | X |  | X | X |  | X | X |
| PC_upto7d_postCOVID \| MinorTherapeutic | 8 | Increased | X | X |  | X |  | X |  |  | X |  | X |  |  |  | X | X |  |  |  |
| PC_upto7d_postCOVID \| 0BH17EZ \| Insertion of Endotracheal Airway into Trachea Via Natural or Artificial Opening | 7 | Increased | X |  |  |  |  | X |  | X | X |  | X |  |  |  | X | X |  |  |  |
| PC_upto7d_postCOVID \| 5A1955Z \| Respiratory Ventilation Greater than 96 Consecutive Hours | 7 | Increased | X |  |  |  |  | X |  | X | X |  | X |  |  |  | X | X |  |  |  |
| IM_upto7d_postCOVID \| Dextrose 10% in Water | 7 | Increased | X | X |  | X |  |  |  |  | X |  | X |  |  |  | X | X |  |  |  |
| IM_upto7d_postCOVID \| fentaNYL | 6 | Increased | X |  |  |  |  |  |  | X | X |  | X |  |  |  | X | X |  |  |  |
| IM_upto7d_postCOVID \| propofol | 6 | Increased | X |  |  |  |  |  |  | X | X |  | X |  |  |  | X | X |  |  |  |
| EP_lengthOfStay | 6 | Increased |  |  | X |  |  |  |  |  |  |  |  | X | X |  |  |  | X | X | X |
| PC_upto7d_postCOVID \| 02HV33Z \| Insertion of Infusion Device into Superior Vena Cava Percutaneous Approach | 6 | Increased |  | X |  |  |  |  |  | X | X |  | X |  |  |  | X | X |  |  |  |
| IM_upto7d_postCOVID \| norepinephrine | 5 | Increased | X |  |  |  |  |  |  | X | X |  | X |  |  |  |  | X |  |  |  |
| IM_7dto14d_postCOVID \| fentaNYL | 5 | Increased | X |  |  |  |  | X |  |  | X |  | X |  |  |  |  | X |  |  |  |
| IM_upto7d_postCOVID \| midazolam | 4 | Increased | X |  |  |  |  |  |  |  | X |  | X |  |  |  |  | X |  |  |  |
| IM_upto7d_postCOVID \| cisatracurium | 4 | Increased | X |  |  |  |  |  |  |  | X |  |  |  |  |  | X | X |  |  |  |
| IM_14dto21d_postCOVID \| fentaNYL | 4 | Increased | X |  |  |  |  |  |  | X |  |  |  |  |  |  | X | X |  |  |  |
| CV_lympho_threshold_flag | 3 | Increased | X |  | X |  |  |  |  |  |  |  | X |  |  |  |  |  |  |  |  |
| IM_upto7d_postCOVID \| furosemide | 3 | Increased | X |  |  |  |  |  |  |  |  |  | X |  |  |  |  | X |  |  |  |
| CV_temp_threshold_flag | 3 | Increased | X |  | X |  |  |  |  |  |  |  |  |  |  |  |  |  |  | X |  |
| VS_>28d_postCOVID \| 3336070 \| O2 Saturation | 3 | Increased |  |  |  |  |  |  | X |  |  | X |  |  |  | X |  |  |  |  |  |
| IM_>28d_postCOVID \| albumin human | 3 | Increased |  | X |  | X |  | X |  |  |  |  |  |  |  |  |  |  |  |  |  |
| IM_>28d_postCOVID \| norepinephrine | 3 | Increased |  |  |  |  |  |  |  | X |  |  | X |  |  |  | X |  |  |  |  |
| IM_upto1m_preCOVID \| norepinephrine | 3 | Increased |  |  | X |  |  |  |  |  |  |  |  |  | X |  |  |  |  |  | X |
| IM_upto1m_preCOVID \| propofol | 3 | Increased |  |  | X |  |  |  |  |  |  |  |  |  | X |  |  |  |  |  | X |
| IM_>28d_postCOVID \| cisatracurium | 3 | Increased |  | X |  |  |  |  |  |  |  |  |  |  |  |  | X | X |  |  |  |
| PC_upto7d_postCOVID \| MinorDiagnostic | 3 | Increased |  | X |  | X |  |  |  |  |  |  |  |  |  |  | X |  |  |  |  |
| LB_>28d_postCOVID \| 1920-8 \| AST SerPl-cCnc | 2 | Increased |  |  |  | X |  |  |  |  |  |  |  |  |  | X |  |  |  |  |  |
| LB_>28d_postCOVID \| 1742-6 \| ALT SerPl-cCnc | 2 | Increased |  |  |  | X |  |  |  |  |  |  |  |  |  | X |  |  |  |  |  |
| DG_>28d_postCOVID \| J80 \| Acute respiratory distress syndrome | 2 | Increased |  |  |  |  |  |  |  | X |  | X |  |  |  |  |  |  |  |  |  |
| IM_upto7d_postCOVID \| albumin human | 2 | Increased |  |  |  |  |  |  |  |  |  |  | X |  |  |  |  | X |  |  |  |
| IM_upto7d_postCOVID \| rocuronium | 2 | Increased |  |  |  |  |  |  |  |  | X |  | X |  |  |  |  |  |  |  |  |
| IM_upto1m_preCOVID \| fentaNYL | 2 | Increased |  |  | X |  |  |  |  |  |  |  |  |  | X |  |  |  |  |  |  |
| IM_14dto21d_postCOVID \| norepinephrine | 2 | Increased |  |  |  |  |  |  |  | X |  |  |  |  |  |  | X |  |  |  |  |
| IM_>28d_postCOVID \| fentaNYL | 2 | Increased |  |  |  |  |  |  |  | X | X |  |  |  |  |  |  |  |  |  |  |
| DG_>28d_postCOVID \| J12.89 \| Other viral pneumonia | 2 | Increased |  |  |  | X |  |  |  | X |  |  |  |  |  |  |  |  |  |  |  |
| IM_>28d_postCOVID \| midazolam | 2 | Increased |  | X |  |  |  |  |  | X |  |  |  |  |  |  |  |  |  |  |  |
| IM_>28d_postCOVID \| Dextrose 5% in Water | 2 | Increased |  |  |  | X |  |  |  | X |  |  |  |  |  |  |  |  |  |  |  |
| IM_21d-28d_postCOVID \| fentaNYL | 2 | Increased |  |  |  |  |  |  |  |  | X |  |  |  |  |  | X |  |  |  |  |
| IM_7dto14d_postCOVID \| Dextrose 10% in Water | 2 | Increased |  | X |  |  |  |  |  |  | X |  |  |  |  |  |  |  |  |  |  |
| LB_>28d_postCOVID \| 61152-5 \| Albumin SerPl BCP-mCnc | 2 | Increased |  |  |  | X |  |  |  |  | X |  |  |  |  |  |  |  |  |  |  |
| IM_upto7d_postCOVID \| insulin regular | 2 | Increased |  | X |  |  |  |  |  |  |  |  |  |  |  |  |  | X |  |  |  |
| DG_>28d_postCOVID \| Z51.5 \| Encounter for palliative care | 2 | Increased |  | X |  | X |  |  |  |  |  |  |  |  |  |  |  |  |  |  |  |
| LB_upto7d_postCOVID \| 48067-3 \| D dimer FEU PPP IA-mCnc | 2 | Increased |  | X |  | X |  |  |  |  |  |  |  |  |  |  |  |  |  |  |  |
| IM_upto1m_preCOVID \| midazolam | 2 | Increased |  |  | X |  |  |  |  |  |  |  |  |  |  |  |  |  |  |  | X |
| IM_upto7d_postCOVID \| Sodium Chloride 0.9% | 1 | Increased | X |  |  |  |  |  |  |  |  |  |  |  |  |  |  |  |  |  |  |
| CV_cough_threshold_flag | 1 | Increased | X |  |  |  |  |  |  |  |  |  |  |  |  |  |  |  |  |  |  |
| IM_upto7d_postCOVID \| vancomycin | 1 | Increased | X |  |  |  |  |  |  |  |  |  |  |  |  |  |  |  |  |  |  |
| IM_upto7d_postCOVID \| acetaminophen | 1 | Increased | X |  |  |  |  |  |  |  |  |  |  |  |  |  |  |  |  |  |  |
| IM_upto1m_preCOVID \| vancomycin | 1 | Increased |  |  |  |  |  |  |  |  |  |  |  |  |  |  |  |  | X |  |  |
| IM_14dto21d_postCOVID \| cisatracurium | 1 | Increased |  |  |  |  |  |  |  |  |  | X |  |  |  |  |  |  |  |  |  |
| IM_7dto14d_postCOVID \| NxStage dialysate | 1 | Increased |  |  |  |  |  |  |  |  |  | X |  |  |  |  |  |  |  |  |  |
| IM_>28d_postCOVID \| Dextrose 10% in Water | 1 | Increased |  |  |  |  |  |  |  |  |  | X |  |  |  |  |  |  |  |  |  |
| IM_14dto21d_postCOVID \| NxStage dialysate | 1 | Increased |  |  |  |  |  |  |  |  |  | X |  |  |  |  |  |  |  |  |  |
| IM_>28d_postCOVID \| phenylephrine | 1 | Increased |  |  |  |  |  |  |  |  |  | X |  |  |  |  |  |  |  |  |  |
| VS_over12m_preCOVID \| 196805942 \| Procalcitonin Lvl. | 1 | Increased |  |  |  |  |  |  |  |  |  | X |  |  |  |  |  |  |  |  |  |
| IM_>28d_postCOVID \| Sodium Chloride 0.9% | 1 | Increased |  |  |  |  |  |  |  |  |  | X |  |  |  |  |  |  |  |  |  |
| IM_7dto14d_postCOVID \| Lactated Ringers Intravenous | 1 | Increased |  |  |  |  |  |  |  |  |  | X |  |  |  |  |  |  |  |  |  |
| LB_>28d_postCOVID \| 6301-6 \| INR PPP | 1 | Increased |  |  |  |  |  |  |  |  |  | X |  |  |  |  |  |  |  |  |  |
| DG_>28d_postCOVID \| R50.9 \| Fever unspecified | 1 | Increased |  |  |  |  |  |  |  |  |  | X |  |  |  |  |  |  |  |  |  |
| PC_upto7d_postCOVID \| MajorDiagnostic | 1 | Increased |  |  |  |  | X |  |  |  |  |  |  |  |  |  |  |  |  |  |  |
| IM_7dto14d_postCOVID \| Sodium Chloride 0.9% | 1 | Increased |  |  |  |  |  | X |  |  |  |  |  |  |  |  |  |  |  |  |  |
| LB_7dto14d_postCOVID \| 61152-5 \| Albumin SerPl BCP-mCnc | 1 | Increased |  |  |  |  |  | X |  |  |  |  |  |  |  |  |  |  |  |  |  |
| IM_upto7d_postCOVID \| magnesium sulfate | 1 | Increased |  |  |  |  |  | X |  |  |  |  |  |  |  |  |  |  |  |  |  |
| LB_>28d_postCOVID \| 742-7 \| Monocytes # Bld Auto | 1 | Increased |  |  |  |  |  | X |  |  |  |  |  |  |  |  |  |  |  |  |  |
| VS_>28d_postCOVID \| 196805942 \| Procalcitonin Lvl. | 1 | Increased |  |  |  |  |  |  | X |  |  |  |  |  |  |  |  |  |  |  |  |
| LB_21d-28d_postCOVID \| 736-9 \| Lymphocytes/leuk NFr Bld Auto | 1 | Increased |  |  |  |  |  |  | X |  |  |  |  |  |  |  |  |  |  |  |  |
| LB_21d-28d_postCOVID \| 731-0 \| Lymphocytes # Bld Auto | 1 | Increased |  |  |  |  |  |  | X |  |  |  |  |  |  |  |  |  |  |  |  |
| IM_upto7d_postCOVID \| dexmedetomidine | 1 | Increased |  |  |  |  |  |  |  |  |  |  | X |  |  |  |  |  |  |  |  |
| DG_>28d_postCOVID \| R65.21 \| Severe sepsis with septic shock | 1 | Increased |  |  |  |  |  |  |  |  |  |  | X |  |  |  |  |  |  |  |  |
| IM_upto7d_postCOVID \| amiodarone | 1 | Increased |  |  |  |  |  |  |  |  |  |  | X |  |  |  |  |  |  |  |  |
| IM_21d-28d_postCOVID \| norepinephrine | 1 | Increased |  |  |  |  |  |  |  | X |  |  |  |  |  |  |  |  |  |  |  |
| IM_upto7d_postCOVID \| vasopressin | 1 | Increased |  |  |  |  |  |  |  | X |  |  |  |  |  |  |  |  |  |  |  |
| IM_14dto21d_postCOVID \| albumin human | 1 | Increased |  |  |  |  |  |  |  | X |  |  |  |  |  |  |  |  |  |  |  |
| IM_upto7d_postCOVID \| ocular lubricant | 1 | Increased |  |  |  |  |  |  |  | X |  |  |  |  |  |  |  |  |  |  |  |
| IM_14dto21d_postCOVID \| ketamine | 1 | Increased |  |  |  |  |  |  |  |  | X |  |  |  |  |  |  |  |  |  |  |
| IM_7dto14d_postCOVID \| potassium chloride | 1 | Increased |  |  |  |  |  |  |  |  | X |  |  |  |  |  |  |  |  |  |  |
| IM_7dto14d_postCOVID \| Dextrose 5% in Water | 1 | Increased |  | X |  |  |  |  |  |  |  |  |  |  |  |  |  |  |  |  |  |
| IM_21d-28d_postCOVID \| cisatracurium | 1 | Increased |  | X |  |  |  |  |  |  |  |  |  |  |  |  |  |  |  |  |  |
| IM_upto7d_postCOVID \| Dextrose 50% in Water | 1 | Increased |  | X |  |  |  |  |  |  |  |  |  |  |  |  |  |  |  |  |  |
| IM_upto7d_postCOVID \| ketamine | 1 | Increased |  | X |  |  |  |  |  |  |  |  |  |  |  |  |  |  |  |  |  |
| IM_7dto14d_postCOVID \| Dextrose 50% in Water | 1 | Increased |  | X |  |  |  |  |  |  |  |  |  |  |  |  |  |  |  |  |  |
| IM_14dto21d_postCOVID \| EPINEPHrine | 1 | Increased |  | X |  |  |  |  |  |  |  |  |  |  |  |  |  |  |  |  |  |
| IM_upto7d_postCOVID \| tocilizumab | 1 | Increased |  | X |  |  |  |  |  |  |  |  |  |  |  |  |  |  |  |  |  |
| IM_>28d_postCOVID \| ketamine | 1 | Increased |  | X |  |  |  |  |  |  |  |  |  |  |  |  |  |  |  |  |  |
| IM_>28d_postCOVID \| sodium bicarbonate | 1 | Increased |  |  |  | X |  |  |  |  |  |  |  |  |  |  |  |  |  |  |  |
| IM_>28d_postCOVID \| amiodarone | 1 | Increased |  |  |  | X |  |  |  |  |  |  |  |  |  |  |  |  |  |  |  |
| LB_14dto21d_postCOVID \| 61152-5 \| Albumin SerPl BCP-mCnc | 1 | Increased |  |  |  | X |  |  |  |  |  |  |  |  |  |  |  |  |  |  |  |
| DG_>28d_postCOVID \| Z66 \| Do not resuscitate | 1 | Increased |  |  |  | X |  |  |  |  |  |  |  |  |  |  |  |  |  |  |  |
| LB_21d-28d_postCOVID \| 61152-5 \| Albumin SerPl BCP-mCnc | 1 | Increased |  |  |  | X |  |  |  |  |  |  |  |  |  |  |  |  |  |  |  |
| IM_>28d_postCOVID \| sodium phosphate | 1 | Increased |  |  |  | X |  |  |  |  |  |  |  |  |  |  |  |  |  |  |  |
| DG_7dto14d_postCOVID \| Z51.5 \| Encounter for palliative care | 1 | Increased |  |  |  | X |  |  |  |  |  |  |  |  |  |  |  |  |  |  |  |
| IM_upto1m_preCOVID \| cisatracurium | 1 | Increased |  |  | X |  |  |  |  |  |  |  |  |  |  |  |  |  |  |  |  |
| IM_upto1m_preCOVID \| succinylcholine | 1 | Increased |  |  | X |  |  |  |  |  |  |  |  |  |  |  |  |  |  |  |  |
| IM_upto1m_preCOVID \| dexmedetomidine | 1 | Increased |  |  | X |  |  |  |  |  |  |  |  |  |  |  |  |  |  |  |  |
| LB_upto1m_preCOVID \| 711-2 \| Eosinophil # Bld Auto | 1 | Increased |  |  | X |  |  |  |  |  |  |  |  |  |  |  |  |  |  |  |  |
| LB_upto1m_preCOVID \| 742-7 \| Monocytes # Bld Auto | 1 | Increased |  |  | X |  |  |  |  |  |  |  |  |  |  |  |  |  |  |  |  |
| LB_upto1m_preCOVID \| 713-8 \| Eosinophil/leuk NFr Bld Auto | 1 | Increased |  |  | X |  |  |  |  |  |  |  |  |  |  |  |  |  |  |  |  |
| IM_upto1m_preCOVID \| tocilizumab | 1 | Increased |  |  | X |  |  |  |  |  |  |  |  |  |  |  |  |  |  |  |  |
| LB_upto7d_postCOVID \| 61152-5 \| Albumin SerPl BCP-mCnc | 1 | Increased |  |  |  |  |  |  |  |  |  |  |  |  |  |  | X |  |  |  |  |
| IM_21d-28d_postCOVID \| midazolam | 1 | Increased |  |  |  |  |  |  |  |  |  |  |  |  |  |  | X |  |  |  |  |
| IM_>28d_postCOVID \| vasopressin | 1 | Increased |  |  |  |  |  |  |  |  |  |  |  |  |  |  | X |  |  |  |  |
| IM_14dto21d_postCOVID \| sodium bicarbonate | 1 | Increased |  |  |  |  |  |  |  |  |  |  |  |  |  |  | X |  |  |  |  |
| IM_upto7d_postCOVID \| calcium gluconate | 1 | Increased |  |  |  |  |  |  |  |  |  |  |  |  |  |  |  | X |  |  |  |
| IM_>28d_postCOVID \| propofol | 1 | Increased |  |  |  |  |  |  |  |  |  |  |  |  |  |  |  | X |  |  |  |
| IM_upto1m_preCOVID \| etomidate | 1 | Increased |  |  |  |  |  |  |  |  |  |  |  |  |  |  |  |  |  |  | X |
| LB_>28d_postCOVID \| 2951-2 \| Sodium SerPl-sCnc | 1 | NA |  |  |  |  |  |  |  |  |  | X |  |  |  |  |  |  |  |  |  |
| LB_upto1m_preCOVID \| 6598-7 \| Troponin T SerPl-mCnc | 1 | NA |  |  |  |  |  |  |  |  |  | X |  |  |  |  |  |  |  |  |  |
| LB_>28d_postCOVID \| 6598-7 \| Troponin T SerPl-mCnc | 1 | NA |  |  |  |  |  |  |  |  |  | X |  |  |  |  |  |  |  |  |  |
| LB_upto7d_postCOVID \| 2341-6 \| Glucose Bld Manual Strip-mCnc | 1 | NA |  |  |  |  |  |  |  |  |  | X |  |  |  |  |  |  |  |  |  |
| VS_>28d_postCOVID \| 3337143 \| Diastolic Blood Pressure NBP | 1 | NA |  |  |  |  | X |  |  |  |  |  |  |  |  |  |  |  |  |  |  |
| VS_>28d_postCOVID \| 3337090 \| Systolic Blood Pressure NBP | 1 | NA |  |  |  |  | X |  |  |  |  |  |  |  |  |  |  |  |  |  |  |
| VS_over12m_preCOVID \| 3357311 \| Abs Lymphocyte Cnt | 1 | NA |  |  | X |  |  |  |  |  |  |  |  |  |  |  |  |  |  |  |  |
