## Supplementary File Table 4 for "Ondansetron use is associated with lower COVID-19 mortality in a Real-World Data network-based analysis"

**Supplemental Table 4**

| **Network Name** | **Feature Name** | **p-value** |
| --- | --- | --- |
| Inpatients age 60+ pre-COVID-19+ PCR Test | EP_lengthOfStay | <1e-10 |
| Inpatients pre-COVID-19+ PCR Test | EP_lengthOfStay | <1e-10 |
| Inpatients Hispanic pre-COVID-19+ PCR Test | EP_lengthOfStay | <1e-10 |
| Inpatients age < 60 pre-COVID-19+ PCR Test | EP_lengthOfStay | 2.00E-10 |
| Inpatients White Non-Hispanic pre-COVID-19+ PCR Test | EP_lengthOfStay | 2.00E-06 |
| Inpatients African American Non-Hispanic pre-COVID-19+ PCR Test | EP_lengthOfStay | 8.00E-05 |
