## Supplementary File Table 5 for "Ondansetron use is associated with lower COVID-19 mortality in a Real-World Data network-based analysis"

|  | **No Ondansetron** | **Ondansetron** | **All** |
| --- | --- | --- | --- |
| **Deceased** | 212 (80.92%) | 50 (19.08%) | 262 |
| **On Ventilator** | 299 (67.34%) | 145 (32.66%) | 444 |
| **Female** | 934 (63.41%) | 539 (36.59%) | 1,473 |
| **Male** | 1,224 (76.07% | 385 (23.93%) | 1609 |
| **White** | 1,168 (69.77%) | 506 (30.23%) | 1674 |
| **Not White** | 953 (70.07%) | 407 (29.93%) | 1360 |
| **NA** | 37 (77.08%) | 11 (22.92%) | 48 |
| **Hispanic** | 835 (67.83%) | 396 (32.17%) | 1231 |
| **Not Hispanic** | 1241 (71.45%) | 496 (28.55%) | 1737 |
| **NA** | 82 (71.93%) | 32 (28.07%) | 114 |
| **18-39** | 239 (61.44%) | 150 (38.56%) | 389 |
| **40-49** | 254 (64.96%) | 137 (35.04%) | 391 |
| **50-59** | 360 (65.69%) | 188 (34.31%) | 548 |
| **60-69** | 466 (71.15%) | 189 (28.85%) | 655 |
| **70+** | 839 (76.34%) | 260 (23.66%) | 1,099 |
| **Heart Failure** | 325 (71.90%) | 127 (28.10%) | 452 |
| **COPD** | 250 (75.76%) | 80 (24.24%) | 330 |
| **Asthma** | 210 (60.34%) | 138 (39.66%) | 348 |
| **Kidney Disease** | 445 (67.63%) | 213 (32.37%) | 658 |
| **Neoplastic Disease** | 118 (63.44%) | 68 (36.56%) | 186 |
| **Dexamethasone** | 391(72.41%) | 149 (27.59%) | 540 |
| **Tocilizumab** | 195 (66.10%) | 100 (33.90%) | 295 |
| **Convalescent plasma** | 217 (67.60%) | 104 (32.40%) | 321 |
| **Remdesivir** | 824 (71.16%) | 334 (28.84%) | 1,158 |
| **Azithromycin** | 820 (67.88%) | 388 (32.12%) | 1,208 |
| **All** | 2,158 (70.02%) | 924 (29.98%) | 3,082 |

**Supplemental Table 5**
