## Supplementary File Table 1 for "Ondansetron use is associated with lower COVID-19 mortality in a Real-World Data network-based analysis"

**Supplemental Table 1.**

| Domain | Logical Table Name | Description |
| --- | --- | --- |
| Subject | Recover_Patient | Patient Cohort table - A collection of unique patient IDs that were tested for the SARS-CoV-2 virus. |
| Interaction | Recover_Encounter | Encounter Cohort table – COVID-19 tested patient encounters for visit types of Emergency, Inpatient, Observation and Outpatient. |
| Diagnosis | Recover_Diagnosis | Diagnosis table - All ICD-10-CM Diagnosis data coded for each Encounter. |
| Diagnosis | Recover_Patient _Problems | Patient Problem table - All SNOMED-CT data coded for each RECOVER-19 Encounter. |
| Procedure | Recover_Procedure | Procedure table - All ICD_10_PCS Procedure data coded for each RECOVER-19 Encounter. |
| Results | Recover_Clinical_Events | Clinical Events table - A specific list of Clinical Event data coded for each RECOVER-19 Encounter. |
| Results | Recover_Lab_Results | Lab Result table - A specific list of LOINC coded lab results for each RECOVER-19 Encounter. |
| Therapy | Recover_Medication_Admin | Medication Administration table - All Medical Administration Recorded events for each RECOVER-19 Encounter. |
| Therapy | Recover_Medication_Home | Medication Home table - Home Medications and Discharged prescribed medications for each RECOVER-19 Encounter. |
