## Supplementary File Table 2 for "Ondansetron use is associated with lower COVID-19 mortality in a Real-World Data network-based analysis"

**Supplemental Table 2**

| Network Name | Feature Name | p-value | OR |
| --- | --- | --- | --- |
| All Patients | CV_cough_threshold_flag | <1e-10 | 21 |
| All Patients | CV_lympho_threshold_flag | <1e-10 | 55 |
| Inpatients pre-COVID-19+ PCR Test | CV_lympho_threshold_flag | <1e-10 | 10 |
| Inpatients age 60+ | CV_lympho_threshold_flag | <1e-10 | 7.2 |
| All Patients | CV_temp_threshold_flag | <1e-10 | 17 |
| Inpatients Hispanic pre-COVID-19+ PCR Test | CV_temp_threshold_flag | 2.00E-08 | 3.6 |
| Inpatients pre-COVID-19+ PCR Test | CV_temp_threshold_flag | <1e-10 | 2.7 |
| Inpatients age 60 to 69 | DG_>28d_postCOVID \| J12.89 \| Other viral pneumonia | 6.00E-06 | 27 |
| Inpatients post-COVID-19+ PCR Test | DG_>28d_postCOVID \| J12.89 \| Other viral pneumonia | <1e-10 | 8.7 |
| Inpatients age < 60 | DG_>28d_postCOVID \| J80 \| Acute respiratory distress syndrome | 7.00E-07 | 9.9 |
| Inpatients age 60 to 69 | DG_>28d_postCOVID \| J80 \| Acute respiratory distress syndrome | 2.00E-04 | 7 |
| Inpatients age < 60 | DG_>28d_postCOVID \| R50.9 \| Fever unspecified | 0.04 | 2.8 |
| Inpatients age 60+ | DG_>28d_postCOVID \| R65.21 \| Severe sepsis with septic shock | <1e-10 | 14 |
| Inpatients | DG_>28d_postCOVID \| Z51.5 \| Encounter for palliative care | <1e-10 | 10 |
| Inpatients post-COVID-19+ PCR Test | DG_>28d_postCOVID \| Z51.5 \| Encounter for palliative care | <1e-10 | 10 |
| Inpatients post-COVID-19+ PCR Test | DG_>28d_postCOVID \| Z66 \| Do not resuscitate | <1e-10 | 7.6 |
| Inpatients post-COVID-19+ PCR Test | DG_7dto14d_postCOVID \| Z51.5 \| Encounter for palliative care | 7.00E-06 | 3.2 |
| All Patients | EP_admittedToICU | <1e-10 | 33 |
| Inpatients age < 60 pre-COVID-19+ PCR Test | EP_admittedToICU | <1e-10 | 12 |
| Inpatients age 60 to 69 | EP_admittedToICU | <1e-10 | 8.1 |
| Inpatients Hispanic | EP_admittedToICU | <1e-10 | 7.4 |
| Inpatients Hispanic pre-COVID-19+ PCR Test | EP_admittedToICU | <1e-10 | 7.4 |
| Inpatients White Non-Hispanic | EP_admittedToICU | <1e-10 | 7.4 |
| Inpatients White Non-Hispanic pre-COVID-19+ PCR Test | EP_admittedToICU | <1e-10 | 7.4 |
| Inpatients pre-COVID-19+ PCR Test | EP_admittedToICU | <1e-10 | 6.5 |
| Inpatients age 60+ | EP_admittedToICU | <1e-10 | 5.3 |
| Inpatients age 60+ pre-COVID-19+ PCR Test | EP_admittedToICU | <1e-10 | 5.3 |
| Inpatients age 70+ | EP_admittedToICU | <1e-10 | 4.5 |
| All Patients | EP_placedOnVentilator | <1e-10 | 170 |
| Inpatients age < 60 | EP_placedOnVentilator | <1e-10 | 170 |
| Inpatients age < 60 pre-COVID-19+ PCR Test | EP_placedOnVentilator | <1e-10 | 170 |
| Inpatients age 40 to 49 | EP_placedOnVentilator | <1e-10 | 85 |
| Inpatients age 60 to 69 | EP_placedOnVentilator | <1e-10 | 73 |
| Inpatients Hispanic | EP_placedOnVentilator | <1e-10 | 38 |
| Inpatients Hispanic pre-COVID-19+ PCR Test | EP_placedOnVentilator | <1e-10 | 38 |
| Inpatients African American Non-Hispanic pre-COVID-19+ PCR Test | EP_placedOnVentilator | <1e-10 | 36 |
| Inpatients | EP_placedOnVentilator | <1e-10 | 33 |
| Inpatients post-COVID-19+ PCR Test | EP_placedOnVentilator | <1e-10 | 33 |
| Inpatients pre-COVID-19+ PCR Test | EP_placedOnVentilator | <1e-10 | 33 |
| Inpatients White Non-Hispanic | EP_placedOnVentilator | <1e-10 | 24 |
| Inpatients White Non-Hispanic pre-COVID-19+ PCR Test | EP_placedOnVentilator | <1e-10 | 24 |
| Inpatients age 60+ | EP_placedOnVentilator | <1e-10 | 23 |
| Inpatients age 60+ pre-COVID-19+ PCR Test | EP_placedOnVentilator | <1e-10 | 23 |
| Inpatients age 70+ | EP_placedOnVentilator | <1e-10 | 19 |
| Inpatients age 40 to 49 | IM_>28d_postCOVID \| albumin human | 0.009 | 18 |
| Inpatients | IM_>28d_postCOVID \| albumin human | <1e-10 | 9.9 |
| Inpatients post-COVID-19+ PCR Test | IM_>28d_postCOVID \| albumin human | <1e-10 | 9.9 |
| Inpatients post-COVID-19+ PCR Test | IM_>28d_postCOVID \| amiodarone - NO MIDLINE IF IV | <1e-10 | 19 |
| Inpatients White Non-Hispanic | IM_>28d_postCOVID \| cisatracurium | 2.00E-10 | 150 |
| Inpatients | IM_>28d_postCOVID \| cisatracurium | <1e-10 | 52 |
| Inpatients Hispanic | IM_>28d_postCOVID \| cisatracurium | <1e-10 | 34 |
| Inpatients age < 60 | IM_>28d_postCOVID \| Dextrose 10% in Water | 5.00E-05 | 7 |
| Inpatients age 60 to 69 | IM_>28d_postCOVID \| Dextrose 5% in Water | 2.00E-04 | 7.3 |
| Inpatients post-COVID-19+ PCR Test | IM_>28d_postCOVID \| Dextrose 5% in Water | <1e-10 | 6 |
| Inpatients age 60 to 69 | IM_>28d_postCOVID \| fentaNYL | 4.00E-08 | 23 |
| Inpatients age 70+ | IM_>28d_postCOVID \| fentaNYL | <1e-10 | 20 |
| Inpatients | IM_>28d_postCOVID \| ketamine | <1e-10 | 13 |
| Inpatients | IM_>28d_postCOVID \| midazolam | <1e-10 | 11 |
| Inpatients age 60 to 69 | IM_>28d_postCOVID \| midazolam | 2.00E-05 | 9.7 |
| Inpatients Hispanic | IM_>28d_postCOVID \| norepinephrine - NO MIDLINE IF IV | <1e-10 | 30 |
| Inpatients age 60 to 69 | IM_>28d_postCOVID \| norepinephrine - NO MIDLINE IF IV | 5.00E-09 | 26 |
| Inpatients age 60+ | IM_>28d_postCOVID \| norepinephrine - NO MIDLINE IF IV | <1e-10 | 25 |
| Inpatients age < 60 | IM_>28d_postCOVID \| phenylephrine - NO MIDLINE IF IV | 0.002 | 7.1 |
| Inpatients White Non-Hispanic | IM_>28d_postCOVID \| propofol - NO MIDLINE IF IV | 6.00E-10 | 68 |
| Inpatients post-COVID-19+ PCR Test | IM_>28d_postCOVID \| sodium bicarbonate - NO MIDLINE IF IV | <1e-10 | 19 |
| Inpatients age < 60 | IM_>28d_postCOVID \| Sodium Chloride 0.9% | 0.003 | 5.4 |
| Inpatients post-COVID-19+ PCR Test | IM_>28d_postCOVID \| sodium phosphate | 1.00E-07 | 6.4 |
| Inpatients Hispanic | IM_>28d_postCOVID \| vasopressin - NO MIDLINE IF IV | <1e-10 | 28 |
| Inpatients age 60 to 69 | IM_14dto21d_postCOVID \| albumin human | 1.00E-06 | 8.4 |
| Inpatients age < 60 | IM_14dto21d_postCOVID \| cisatracurium | <1e-10 | 13 |
| Inpatients | IM_14dto21d_postCOVID \| EPINEPHrine - NO MIDLINE IF IV | <1e-10 | 23 |
| All Patients | IM_14dto21d_postCOVID \| fentaNYL | <1e-10 | 40 |
| Inpatients age 60 to 69 | IM_14dto21d_postCOVID \| fentaNYL | <1e-10 | 28 |
| Inpatients White Non-Hispanic | IM_14dto21d_postCOVID \| fentaNYL | <1e-10 | 25 |
| Inpatients Hispanic | IM_14dto21d_postCOVID \| fentaNYL | <1e-10 | 12 |
| Inpatients age 70+ | IM_14dto21d_postCOVID \| ketamine | 6.00E-07 | 8.8 |
| Inpatients age 60 to 69 | IM_14dto21d_postCOVID \| norepinephrine - NO MIDLINE IF IV | 1.00E-10 | 13 |
| Inpatients Hispanic | IM_14dto21d_postCOVID \| norepinephrine - NO MIDLINE IF IV | <1e-10 | 12 |
| Inpatients age < 60 | IM_14dto21d_postCOVID \| NxStage dialysate | 0.002 | 8.7 |
| Inpatients Hispanic | IM_14dto21d_postCOVID \| sodium bicarbonate - NO MIDLINE IF IV | <1e-10 | 45 |
| Inpatients | IM_21d-28d_postCOVID \| cisatracurium | <1e-10 | 16 |
| Inpatients Hispanic | IM_21d-28d_postCOVID \| fentaNYL | <1e-10 | 18 |
| Inpatients age 70+ | IM_21d-28d_postCOVID \| fentaNYL | <1e-10 | 15 |
| Inpatients Hispanic | IM_21d-28d_postCOVID \| midazolam | <1e-10 | 16 |
| Inpatients age 60 to 69 | IM_21d-28d_postCOVID \| norepinephrine - NO MIDLINE IF IV | <1e-10 | 29 |
| Inpatients | IM_7dto14d_postCOVID \| Dextrose 10% in Water | <1e-10 | 5.1 |
| Inpatients age 70+ | IM_7dto14d_postCOVID \| Dextrose 10% in Water | <1e-10 | 4.9 |
| Inpatients | IM_7dto14d_postCOVID \| Dextrose 5% in Water | <1e-10 | 9.1 |
| Inpatients | IM_7dto14d_postCOVID \| Dextrose 50% in Water | <1e-10 | 5.7 |
| All Patients | IM_7dto14d_postCOVID \| fentaNYL | <1e-10 | 31 |
| Inpatients age 40 to 49 | IM_7dto14d_postCOVID \| fentaNYL | 6.00E-06 | 16 |
| Inpatients White Non-Hispanic | IM_7dto14d_postCOVID \| fentaNYL | <1e-10 | 16 |
| Inpatients age 70+ | IM_7dto14d_postCOVID \| fentaNYL | <1e-10 | 14 |
| Inpatients age 60+ | IM_7dto14d_postCOVID \| fentaNYL | <1e-10 | 13 |
| Inpatients age < 60 | IM_7dto14d_postCOVID \| Lactated Ringers Intravenous | 0.009 | 3.4 |
| Inpatients age < 60 | IM_7dto14d_postCOVID \| NxStage dialysate | 6.00E-08 | 25 |
| Inpatients age 70+ | IM_7dto14d_postCOVID \| potassium chloride | 6.00E-06 | 2.9 |
| Inpatients age 40 to 49 | IM_7dto14d_postCOVID \| Sodium Chloride 0.9% | 0.002 | 5.9 |
| Inpatients pre-COVID-19+ PCR Test | IM_upto1m_preCOVID \| cisatracurium | 4.00E-09 | 6.1 |
| Inpatients pre-COVID-19+ PCR Test | IM_upto1m_preCOVID \| dexmedetomidine | 6.00E-04 | 3.5 |
| Inpatients White Non-Hispanic pre-COVID-19+ PCR Test | IM_upto1m_preCOVID \| etomidate | 0.01 | 3.6 |
| Inpatients pre-COVID-19+ PCR Test | IM_upto1m_preCOVID \| fentaNYL | <1e-10 | 5.8 |
| Inpatients age 60+ pre-COVID-19+ PCR Test | IM_upto1m_preCOVID \| fentaNYL | 3.00E-09 | 4.1 |
| Inpatients pre-COVID-19+ PCR Test | IM_upto1m_preCOVID \| midazolam | <1e-10 | 4.8 |
| Inpatients White Non-Hispanic pre-COVID-19+ PCR Test | IM_upto1m_preCOVID \| midazolam | 0.001 | 3.8 |
| Inpatients pre-COVID-19+ PCR Test | IM_upto1m_preCOVID \| norepinephrine - NO MIDLINE IF IV | <1e-10 | 8.2 |
| Inpatients White Non-Hispanic pre-COVID-19+ PCR Test | IM_upto1m_preCOVID \| norepinephrine - NO MIDLINE IF IV | 7.00E-06 | 5.8 |
| Inpatients age 60+ pre-COVID-19+ PCR Test | IM_upto1m_preCOVID \| norepinephrine - NO MIDLINE IF IV | 3.00E-09 | 5 |
| Inpatients pre-COVID-19+ PCR Test | IM_upto1m_preCOVID \| propofol - NO MIDLINE IF IV | <1e-10 | 6 |
| Inpatients White Non-Hispanic pre-COVID-19+ PCR Test | IM_upto1m_preCOVID \| propofol - NO MIDLINE IF IV | 5.00E-04 | 4.2 |
| Inpatients age 60+ pre-COVID-19+ PCR Test | IM_upto1m_preCOVID \| propofol - NO MIDLINE IF IV | 9.00E-07 | 3.8 |
| Inpatients pre-COVID-19+ PCR Test | IM_upto1m_preCOVID \| succinylcholine | 2.00E-04 | 4.8 |
| Inpatients pre-COVID-19+ PCR Test | IM_upto1m_preCOVID \| tocilizumab | 0.03 | 2.7 |
| Inpatients African American Non-Hispanic pre-COVID-19+ PCR Test | IM_upto1m_preCOVID \| vancomycin | 0.02 | 2.4 |
| All Patients | IM_upto7d_postCOVID \| acetaminophen | <1e-10 | 11 |
| Inpatients White Non-Hispanic | IM_upto7d_postCOVID \| albumin human | 4.00E-10 | 11 |
| Inpatients age 60+ | IM_upto7d_postCOVID \| albumin human | <1e-10 | 7.5 |
| Inpatients age 60+ | IM_upto7d_postCOVID \| amiodarone - NO MIDLINE IF IV | <1e-10 | 9.6 |
| Inpatients White Non-Hispanic | IM_upto7d_postCOVID \| calcium gluconate | 4.00E-10 | 20 |
| All Patients | IM_upto7d_postCOVID \| cisatracurium | <1e-10 | 55 |
| Inpatients White Non-Hispanic | IM_upto7d_postCOVID \| cisatracurium | <1e-10 | 12 |
| Inpatients age 70+ | IM_upto7d_postCOVID \| cisatracurium | <1e-10 | 11 |
| Inpatients Hispanic | IM_upto7d_postCOVID \| cisatracurium | <1e-10 | 7.7 |
| Inpatients age 60+ | IM_upto7d_postCOVID \| dexmedetomidine | <1e-10 | 5.8 |
| All Patients | IM_upto7d_postCOVID \| Dextrose 10% in Water | <1e-10 | 36 |
| Inpatients | IM_upto7d_postCOVID \| Dextrose 10% in Water | <1e-10 | 6.7 |
| Inpatients post-COVID-19+ PCR Test | IM_upto7d_postCOVID \| Dextrose 10% in Water | <1e-10 | 6.7 |
| Inpatients White Non-Hispanic | IM_upto7d_postCOVID \| Dextrose 10% in Water | <1e-10 | 5.8 |
| Inpatients Hispanic | IM_upto7d_postCOVID \| Dextrose 10% in Water | <1e-10 | 5.5 |
| Inpatients age 60+ | IM_upto7d_postCOVID \| Dextrose 10% in Water | <1e-10 | 4.3 |
| Inpatients age 70+ | IM_upto7d_postCOVID \| Dextrose 10% in Water | <1e-10 | 3.5 |
| Inpatients | IM_upto7d_postCOVID \| Dextrose 50% in Water | <1e-10 | 5.4 |
| All Patients | IM_upto7d_postCOVID \| fentaNYL | <1e-10 | 60 |
| Inpatients age 60 to 69 | IM_upto7d_postCOVID \| fentaNYL | <1e-10 | 14 |
| Inpatients White Non-Hispanic | IM_upto7d_postCOVID \| fentaNYL | <1e-10 | 13 |
| Inpatients age 60+ | IM_upto7d_postCOVID \| fentaNYL | <1e-10 | 11 |
| Inpatients age 70+ | IM_upto7d_postCOVID \| fentaNYL | <1e-10 | 10 |
| Inpatients Hispanic | IM_upto7d_postCOVID \| fentaNYL | <1e-10 | 7.6 |
| All Patients | IM_upto7d_postCOVID \| furosemide | <1e-10 | 33 |
| Inpatients White Non-Hispanic | IM_upto7d_postCOVID \| furosemide | <1e-10 | 8 |
| Inpatients age 60+ | IM_upto7d_postCOVID \| furosemide | <1e-10 | 4.8 |
| Inpatients White Non-Hispanic | IM_upto7d_postCOVID \| insulin regular | <1e-10 | 10 |
| Inpatients | IM_upto7d_postCOVID \| insulin regular | <1e-10 | 6.2 |
| Inpatients | IM_upto7d_postCOVID \| ketamine | <1e-10 | 7 |
| Inpatients age 40 to 49 | IM_upto7d_postCOVID \| magnesium sulfate | 0.02 | 5.2 |
| All Patients | IM_upto7d_postCOVID \| midazolam | <1e-10 | 56 |
| Inpatients White Non-Hispanic | IM_upto7d_postCOVID \| midazolam | <1e-10 | 13 |
| Inpatients age 70+ | IM_upto7d_postCOVID \| midazolam | <1e-10 | 9.1 |
| Inpatients age 60+ | IM_upto7d_postCOVID \| midazolam | <1e-10 | 8.9 |
| All Patients | IM_upto7d_postCOVID \| norepinephrine - NO MIDLINE IF IV | <1e-10 | 76 |
| Inpatients White Non-Hispanic | IM_upto7d_postCOVID \| norepinephrine - NO MIDLINE IF IV | <1e-10 | 20 |
| Inpatients age 60 to 69 | IM_upto7d_postCOVID \| norepinephrine - NO MIDLINE IF IV | <1e-10 | 14 |
| Inpatients age 60+ | IM_upto7d_postCOVID \| norepinephrine - NO MIDLINE IF IV | <1e-10 | 11 |
| Inpatients age 70+ | IM_upto7d_postCOVID \| norepinephrine - NO MIDLINE IF IV | <1e-10 | 10 |
| Inpatients age 60 to 69 | IM_upto7d_postCOVID \| ocular lubricant | 1.00E-04 | 8.8 |
| All Patients | IM_upto7d_postCOVID \| propofol - NO MIDLINE IF IV | <1e-10 | 69 |
| Inpatients age 60 to 69 | IM_upto7d_postCOVID \| propofol - NO MIDLINE IF IV | <1e-10 | 16 |
| Inpatients White Non-Hispanic | IM_upto7d_postCOVID \| propofol - NO MIDLINE IF IV | <1e-10 | 14 |
| Inpatients age 60+ | IM_upto7d_postCOVID \| propofol - NO MIDLINE IF IV | <1e-10 | 11 |
| Inpatients age 70+ | IM_upto7d_postCOVID \| propofol - NO MIDLINE IF IV | <1e-10 | 11 |
| Inpatients Hispanic | IM_upto7d_postCOVID \| propofol - NO MIDLINE IF IV | <1e-10 | 9 |
| Inpatients age 70+ | IM_upto7d_postCOVID \| rocuronium | 3.00E-09 | 6.5 |
| Inpatients age 60+ | IM_upto7d_postCOVID \| rocuronium | <1e-10 | 6.5 |
| All Patients | IM_upto7d_postCOVID \| Sodium Chloride 0.9% | <1e-10 | 38 |
| Inpatients | IM_upto7d_postCOVID \| tocilizumab | <1e-10 | 3.9 |
| All Patients | IM_upto7d_postCOVID \| vancomycin | <1e-10 | 22 |
| Inpatients age 60 to 69 | IM_upto7d_postCOVID \| vasopressin - NO MIDLINE IF IV | 2.00E-10 | 31 |
| Inpatients African American Non-Hispanic | LB_>28d_postCOVID \| 1742-6 \| ALT SerPl-cCnc | 0.002 | 9.9 |
| Inpatients post-COVID-19+ PCR Test | LB_>28d_postCOVID \| 1742-6 \| ALT SerPl-cCnc | 3.00E-06 | 4 |
| Inpatients African American Non-Hispanic | LB_>28d_postCOVID \| 1920-8 \| AST SerPl-cCnc | 7.00E-05 | 18 |
| Inpatients post-COVID-19+ PCR Test | LB_>28d_postCOVID \| 1920-8 \| AST SerPl-cCnc | 7.00E-10 | 6.1 |
| Inpatients age 70+ | LB_>28d_postCOVID \| 61152-5 \| Albumin SerPl BCP-mCnc | 2.00E-07 | 12 |
| Inpatients post-COVID-19+ PCR Test | LB_>28d_postCOVID \| 61152-5 \| Albumin SerPl BCP-mCnc | <1e-10 | 7.6 |
| Inpatients age < 60 | LB_>28d_postCOVID \| 6301-6 \| INR PPP | 0.03 | 2.7 |
| Inpatients age 40 to 49 | LB_>28d_postCOVID \| 742-7 \| Monocytes # Bld Auto | 0.03 | 10 |
| Inpatients post-COVID-19+ PCR Test | LB_14dto21d_postCOVID \| 61152-5 \| Albumin SerPl BCP-mCnc | <1e-10 | 5.3 |
| Inpatients post-COVID-19+ PCR Test | LB_21d-28d_postCOVID \| 61152-5 \| Albumin SerPl BCP-mCnc | 2.00E-09 | 6.6 |
| Inpatients age 50 to 59 | LB_21d-28d_postCOVID \| 731-0 \| Lymphocytes # Bld Auto | 0.04 | 4.1 |
| Inpatients age 50 to 59 | LB_21d-28d_postCOVID \| 736-9 \| Lymphocytes/leuk NFr Bld Auto | 0.004 | Inf |
| Inpatients age 40 to 49 | LB_7dto14d_postCOVID \| 61152-5 \| Albumin SerPl BCP-mCnc | 0.005 | 13 |
| Inpatients pre-COVID-19+ PCR Test | LB_upto1m_preCOVID \| 711-2 \| Eosinophil # Bld Auto | 0.004 | 3.3 |
| Inpatients pre-COVID-19+ PCR Test | LB_upto1m_preCOVID \| 713-8 \| Eosinophil/leuk NFr Bld Auto | 0.03 | 2.4 |
| Inpatients pre-COVID-19+ PCR Test | LB_upto1m_preCOVID \| 742-7 \| Monocytes # Bld Auto | 0.01 | 1.7 |
| Inpatients | LB_upto7d_postCOVID \| 48067-3 \| D dimer FEU PPP IA-mCnc | <1e-10 | 4.1 |
| Inpatients post-COVID-19+ PCR Test | LB_upto7d_postCOVID \| 48067-3 \| D dimer FEU PPP IA-mCnc | <1e-10 | 4.1 |
| Inpatients Hispanic | LB_upto7d_postCOVID \| 61152-5 \| Albumin SerPl BCP-mCnc | <1e-10 | 5.1 |
| Inpatients Hispanic | PC_upto7d_postCOVID \| 02HV33Z \| Insertion of Infusion Device into Superior Vena Cava Percutaneous Approach | <1e-10 | 12 |
| Inpatients age 70+ | PC_upto7d_postCOVID \| 02HV33Z \| Insertion of Infusion Device into Superior Vena Cava Percutaneous Approach | <1e-10 | 11 |
| Inpatients White Non-Hispanic | PC_upto7d_postCOVID \| 02HV33Z \| Insertion of Infusion Device into Superior Vena Cava Percutaneous Approach | 4.00E-10 | 10 |
| Inpatients | PC_upto7d_postCOVID \| 02HV33Z \| Insertion of Infusion Device into Superior Vena Cava Percutaneous Approach | <1e-10 | 10 |
| Inpatients age 60+ | PC_upto7d_postCOVID \| 02HV33Z \| Insertion of Infusion Device into Superior Vena Cava Percutaneous Approach | <1e-10 | 8.6 |
| Inpatients age 60 to 69 | PC_upto7d_postCOVID \| 02HV33Z \| Insertion of Infusion Device into Superior Vena Cava Percutaneous Approach | 7.00E-09 | 8.2 |
| All Patients | PC_upto7d_postCOVID \| 0BH17EZ \| Insertion of Endotracheal Airway into Trachea Via Natural or Artificial Opening | <1e-10 | 130 |
| Inpatients age 40 to 49 | PC_upto7d_postCOVID \| 0BH17EZ \| Insertion of Endotracheal Airway into Trachea Via Natural or Artificial Opening | <1e-10 | 120 |
| Inpatients age 60 to 69 | PC_upto7d_postCOVID \| 0BH17EZ \| Insertion of Endotracheal Airway into Trachea Via Natural or Artificial Opening | <1e-10 | 45 |
| Inpatients Hispanic | PC_upto7d_postCOVID \| 0BH17EZ \| Insertion of Endotracheal Airway into Trachea Via Natural or Artificial Opening | <1e-10 | 37 |
| Inpatients age 60+ | PC_upto7d_postCOVID \| 0BH17EZ \| Insertion of Endotracheal Airway into Trachea Via Natural or Artificial Opening | <1e-10 | 19 |
| Inpatients White Non-Hispanic | PC_upto7d_postCOVID \| 0BH17EZ \| Insertion of Endotracheal Airway into Trachea Via Natural or Artificial Opening | <1e-10 | 16 |
| Inpatients age 70+ | PC_upto7d_postCOVID \| 0BH17EZ \| Insertion of Endotracheal Airway into Trachea Via Natural or Artificial Opening | <1e-10 | 15 |
| All Patients | PC_upto7d_postCOVID \| 5A1955Z \| Respiratory Ventilation Greater than 96 Consecutive Hours | <1e-10 | 94 |
| Inpatients age 40 to 49 | PC_upto7d_postCOVID \| 5A1955Z \| Respiratory Ventilation Greater than 96 Consecutive Hours | 7.00E-10 | 39 |
| Inpatients age 60 to 69 | PC_upto7d_postCOVID \| 5A1955Z \| Respiratory Ventilation Greater than 96 Consecutive Hours | <1e-10 | 20 |
| Inpatients Hispanic | PC_upto7d_postCOVID \| 5A1955Z \| Respiratory Ventilation Greater than 96 Consecutive Hours | <1e-10 | 20 |
| Inpatients White Non-Hispanic | PC_upto7d_postCOVID \| 5A1955Z \| Respiratory Ventilation Greater than 96 Consecutive Hours | <1e-10 | 18 |
| Inpatients age 60+ | PC_upto7d_postCOVID \| 5A1955Z \| Respiratory Ventilation Greater than 96 Consecutive Hours | <1e-10 | 13 |
| Inpatients age 70+ | PC_upto7d_postCOVID \| 5A1955Z \| Respiratory Ventilation Greater than 96 Consecutive Hours | <1e-10 | 13 |
| Inpatients age 18 to 39 | PC_upto7d_postCOVID \| MajorDiagnostic | 0.01 | 9.4 |
| Inpatients Hispanic | PC_upto7d_postCOVID \| MinorDiagnostic | <1e-10 | 6.3 |
| Inpatients | PC_upto7d_postCOVID \| MinorDiagnostic | <1e-10 | 3.7 |
| Inpatients post-COVID-19+ PCR Test | PC_upto7d_postCOVID \| MinorDiagnostic | <1e-10 | 3.7 |
| All Patients | PC_upto7d_postCOVID \| MinorTherapeutic | <1e-10 | 62 |
| Inpatients age 40 to 49 | PC_upto7d_postCOVID \| MinorTherapeutic | 1.00E-08 | 29 |
| Inpatients Hispanic | PC_upto7d_postCOVID \| MinorTherapeutic | <1e-10 | 16 |
| Inpatients | PC_upto7d_postCOVID \| MinorTherapeutic | <1e-10 | 13 |
| Inpatients post-COVID-19+ PCR Test | PC_upto7d_postCOVID \| MinorTherapeutic | <1e-10 | 13 |
| Inpatients age 60+ | PC_upto7d_postCOVID \| MinorTherapeutic | <1e-10 | 9.3 |
| Inpatients White Non-Hispanic | PC_upto7d_postCOVID \| MinorTherapeutic | <1e-10 | 9.3 |
| Inpatients age 70+ | PC_upto7d_postCOVID \| MinorTherapeutic | <1e-10 | 6.9 |
| Inpatients age 50 to 59 | VS_>28d_postCOVID \| 196805942 \| Procalcitonin Lvl. | 0.002 | 3.9 |
| Inpatients age 50 to 59 | VS_>28d_postCOVID \| 3336070 \| O2 Saturation | 0.02 | 20 |
| Inpatients age < 60 | VS_>28d_postCOVID \| 3336070 \| O2 Saturation | 0.03 | 11 |
| Inpatients African American Non-Hispanic | VS_>28d_postCOVID \| 3336070 \| O2 Saturation | 0.009 | 10 |
| Inpatients age < 60 | VS_over12m_preCOVID \| 196805942 \| Procalcitonin Lvl. | 0.003 | 11 |
